# Wearable tracking of walking and non-walking as progression markers in early Parkinson’s disease

**DOI:** 10.1101/2025.08.19.25333986

**Authors:** King Chung Ho, Siying Li, Claudia Serrano Amenos, Nathan Kowahl, Erin Rainaldi, Chen Chen, Bastiaan R. Bloem, Laurie H. Sanders, Ludy C. Shih, Andrew Siderowf, William J. Marks, Ritu Kapur, Luc J.W. Evers, Sooyoon Shin

## Abstract

**IMPORTANCE:** Wearable-based measures of walking (as proxy for physical activity) may quantify disease progression and modification thereof in early-stage Parkinson’s disease (PD).

**OBJECTIVES:** Establishing the validity of digital measures of walking and non-walking in PD.

**DESIGN:** Retrospective longitudinal analyses of data from cohorts within 3 larger studies, consisting of wearable sensor, demographic, and clinical data collected during 2017-2023, with 1-2 year follow up.

**SETTING:** Three independent multicenter cohort studies.

**PARTICIPANTS:** People with PD, and age/sex matched non-PD cohort.

**EXPOSURES:** None.

**MAIN OUTCOMES AND MEASURES:** Digital measures’ test-retest reliability, analyzed using intraclass correlation coefficients across consecutive monthly-aggregated data. Digital measures’ sensitivity: ability to detect within-participant changes, analyzed over 24 months using linear mixed-effect models, and analyzed as effect-size changes-from-baseline comparing 1- and 2-year longitudinal Cohen’s-d (mean and 95% CIs) vs conventional clinical endpoints. Analyses replicated in two independent PD cohorts (internal validation and external evaluation). Compared within-participant changes between PD and non-PD cohorts using linear mixed-effect model slopes.

**RESULTS:** We analyzed 57 digital measures (51 individual, 6 composite) in a development cohort (N=171), selecting 32 (26 individual, 6 composite) for further study based on their sensitivity and test-retest reliability. During internal validation (N=101), 20 measures could detect statistically significant within-participant changes and 7 showed larger 2-year effect-size changes than conventional clinical measures; non-walking bout (NWB) duration (12.4% yearly change; 2-year Cohen’s-d 0.623 [95% CI: 0.461,0.811]) and 95th percentile of NWB duration (17.1% yearly change; 2-year Cohen’s-d, 0.623 [95% CI: 0.461,0.811]) performed best. Measures could detect significant and persisting changes from baseline at 10 months. During external evaluation (N=67), 15 measures could detect statistically significant within-participant changes and 12 showed larger 1-year effect-size changes than conventional clinical measures; 12 measures showed significantly greater change in people with PD than in matched non-PD individuals (N=171).

**CONCLUSION AND RELEVANCE:** Internal validation and external evaluation of 32 digital measures that quantified walking and non-walking behaviors in patients with early-stage PD showed that they could have greater sensitivity to detect longitudinal changes than conventional measures, and that these changes were disease-specific (e.g., separate from aging), making them candidates for disease-specific progression markers.

**Key Points:** *Question:* Can wearable sensor-based digital measures of physical activity and mobility serve as markers of disease progression in early-stage Parkinson’s disease (PD)?

*Findings:* In two independent longitudinal cohorts of people with PD, digital measures detected statistically-significant changes in walking and non-walking behaviors after 1 and 2 years of follow-up; additionally, a comparison between people with and without PD (from a third cohort) showed that these changes were disease-specific. Compared with MDS-UPDRS-based conventional metrics, measures of non-walking behavior showed greater effect size (such as mean non-walking bout duration, with an annual increase of 12.4% and a 2-year Cohen’s-d of 0.623).

*Meaning:* Wearable sensor-based digital measures can detect and quantify disease-specific changes in walking and non-walking behaviors over time in people with early-stage PD.

## Introduction

Mobility is a meaningful aspect of health for people with Parkinson’s disease (PD), from early stage and throughout the disease course.^1,2^ Function degradation in people with PD can manifest as walking-behavior changes,^2–8^ which may be used to monitor disease progression.

Conventional clinical assessments may be inadequate to capture functional impairments in early-stage PD progression, because of their subtlety and heterogeneity.^9–12^ Insufficiently sensitive disease-progression markers may impede clinical research in early-stage PD, where development of disease-modifying interventions would ideally take place.^13,14^

Wearable sensor-based digital measures can provide sensitive, granular, objective and real-world assessments of mobility, addressing limitations of conventional clinical measures for neurological conditions.^10,15–21^ However, the adoption of digital measures in clinical research must be rigorous.^22^ Investigators need to determine the sensitivity of measures to quantify disease progression and need to validate them, to ensure their accuracy in reflecting clinically relevant and meaningful aspects of health.

Earlier studies have evaluated digital measures during daily-life monitoring of patients with PD. However, these studies lacked appropriate-sized cohorts, long-term follow-up, replication across independent datasets, or inclusion of digital measures capturing comprehensive aspects of walking and non-walking behaviors.^20,23–33^ This study had the following objectives: to apply a previously developed suite of digital measures of walking and non-walking behaviors^34,35^ in a cohort of individuals with early-stage PD and evaluate their reliability and sensitivity to detect within-subject change compared with conventional clinical measures; to replicate those analyses (assessing the validity of those measures) in a second, non-overlapping, independent cohort; and to evaluate their specificity to disease changes in a third, independent cohort, compared to a demographically-matched cohort without PD.

## Methods

### Study cohorts

Our study sub-cohorts originated from 3 previously reported studies^36–38^ (Suppl Fig 1). These studies complied with the Declaration of Helsinki and good clinical practice (GCP) standards, and all their participants provided informed consent.

The Personalized Parkinson Project (PPP)^36^ is a longitudinal cohort study of people with PD. We selected those with early-stage PD, defined as ≤4 years since diagnosis at enrollment.^39^ Participants wore the wrist sensor device (Verily Study Watch) from enrollment, for 2 years. We split the dataset into development (PPP-development) and internal-validation (PPP-test) cohorts at the participant level (split 60:40 without participant overlap, with comparable distributions of their Movement Disorder Society revision of the Unified Parkinson’s Disease Rating Scale (MDS-UPDRS)-Part III scores). The PPP complied with the Dutch Personal Data Protection Act, and the European General Data Protection Regulation. The Commissie Mensgebonden Onderzoek Region Arnhem Nijmegen (reference number 2016–2934; NL59694.091.17) approved the study protocol and communication materials.

The Parkinson’s Progression Markers Initiative (PPMI)^37,40^ is a longitudinal study including participants with and without PD. Participants in the US wore wrist sensor devices starting (on average) at 55 months after enrolment, and for up to 2 years afterwards. Eligible participants for this study were those with PD diagnosis and any sensor data from the Verily Study Watch (Suppl. Methods). The PPMI was approved by the local ethics committees of the participating sites.

The Project Baseline Health Study (PBHS)^38^ was a longitudinal cohort study of participants without diagnosis-specific selection. Eligible participants were those without PD or associated risk factors (self- or family-related; Suppl. Methods) and with data from the wrist sensor device for up to 56 months. A non-PD cohort was constructed by random sampling to match the PPMI cohort (up to 3 matches), based on age (± 5 years), gender, and month 0 of wear time (within 15 months). A central institutional review board and those of all participating institutions (Duke University, Stanford University, California Health and Longevity Institute) approved the study.

### Study Procedures

The PPP, PPMI and PBHS have been described elsewhere^36–38^ (Suppl Fig 1). The relevant procedures for our analyses were as follows:

Participants were evaluated at baseline and yearly in the PPP and PPMI studies, including: MDS-UPDRS, Schwab & England Activities of Daily Living (ADL) scale, Hoehn and Yahr (H&Y) scale, and Parkinson’s Disease Questionnaire (PDQ)-39 (only in PPP).

PBHS participants were evaluated at baseline for self-reported medical history and general demographic characteristics, and had multi-assessment annual visits (Suppl. Fig.1).

Overall, participants in the 3 studies were instructed to wear the device throughout the day and night (except when charging). During the first year of the PBHS, participants were instructed to wear it for 10 hours (daytime); those instructions were modified after the first year.

### Device

The Verily Study Watch (Verily Life Sciences, Dallas, Texas) is a wrist-worn device with sensors to continuously collect tri-axial accelerometer and gyroscope data (at a sample rate of 100 Hz for PD cohorts and 30 Hz for non-PD cohort), along with photoplethysmogram (PPG) data (30 Hz). The device and measures in this study have been previously described.^34,35,41^

### Data Selection, Availability and Processing

Based on accelerometer data, we estimated step counts in 10-second epochs, using a previously described algorithm.^34,35^ We filtered analyzable epochs in each cohort (Suppl. Methods), to ensure focus on days with sufficient daily wear time to include all physical activity during waking time and exclude sleep time.

### Digital Measures

Epochs were classified as walking- (≥6 steps per epoch) or non-walking. From this, we derived a suite of measures quantifying and characterizing walking or non-walking behaviors (Suppl. Table 1).

#### Individual Measures

Individual measures in this study (largely described previously)^34,35^ were derived from step counts, ambulatory time, cadence (number of steps per second, during walking periods), and bouts (defined as a series of contiguous epochs with similar behavior, either lasting for ≥30 seconds [ie, at least 3 epochs] for walking bouts [WB], or ≥10 seconds [ie, at least 1 epoch] for non-walking bouts [NWB]). The actual measures were reported as daily aggregates for individual participants (e.g., daily mean WB duration) (Suppl. Table 1).

#### Composite Measures

Composite measures (CMs) were linear combinations of individual measures (Suppl. Table 1). Based on results from the PPP-development cohort, we selected individual measures as input features to algorithms that learn the weights of these linear combinations; we picked individual measures that (a) were sensitive to symptom progression (showing statistically significant longitudinal measure changes and in a direction consistent with disease progression), and (b) showed good test-retest reliability month-to-month (intraclass correlation coefficient [ICC] averaged over 1 year ≥0.75). Measures with high measure-to-measure correlations and low measure-to-reference-clinical-score correlations^30^ in the PPP-development analysis were removed. For learning of weights, we used supervised learning (reference-based using clinician-reported scores [CROs] and patient-reported scores [PROs] as labels) and unsupervised approaches (reference-free, to mitigate potential limitations of the training-labels, such as subjectivity or low reliability). The supervised approach in this study was the Elasticnet regressor with the clinical labels: total MDS-UPDRS Part III (excluding tremor, facial expression, and speech), MDS-UPDRS Part II + III, MDS-UPDRS Part III gait and posture score, MDS-UPDRS Parts II+III gait-only score, Schwab and England ADL score, PDQ-39, and PDQ-39 mobility score. We used linear principal component analysis (PCA) as the unsupervised approach (Suppl. Methods).

##### Statistical analyses (Suppl. Table 2)

The criteria to evaluate measures throughout our study were: (a) test-retest reliability, (b) sensitivity to symptom progression, and (c) specificity of the observed changes to PD diagnosis.

The analysis involved three steps: (1) selection of measures based on the PPP-development cohort, based on their reliability and sensitivity to change (the exact criteria are described below), (2) internal validation on the PPP-test cohort, by examining the reliability and sensitivity to change, and (3) external evaluation, replicating the reliability and sensitivity analyses using the PPMI-PD cohort, and examining the specificity of the observed progression to PD using non-PD participants from the PBHS cohort.

#### Test-retest Reliability

We calculated the ICC of monthly-aggregated measure values, generated in consecutive, adjacent and non-overlapping months, and averaged over 1 year. ‘Good test-retest reliability’ was defined as ICC≥0.75.

Measures with ICC<0.75 in the PPP-development cohort were excluded from subsequent investigation. We calculated measures’ ICCs for PPP-test and PPMI-PD cohorts.

#### Sensitivity to Change

Sensitivity was defined as the ability to detect disease progression via within-participant changes (up to 24 months) assessed with linear mixed-effect models (Suppl. Methods), adjusted for time-invariant (age-group, gender, year since diagnosis) and time-variant effects (seasonality and COVID-lockdowns). A measure was determined sensitive if it showed statistically significant changes. Statistical significance was determined based on adjusted p-values, computed using the Benjamini-Hochberg (BH) procedure^42^ to control the false discovery rate (FDR) at alpha-level of 0.05.

Measure sensitivity analyzed in the PPP-development cohort enabled selection of measures for subsequent investigation. The primary analyses examined the sensitivity of the selected measures in the PPP-test cohort. Exploratory sensitivity analyses included additional adjustment for time-varying Levodopa equivalent daily dose (LEDD; log2 transformed), to evaluate any potential impact of concurrent medication.

As secondary analyses, we characterized within-participant changes at 1- and 2-years from baseline in the PPP-test cohort using longitudinal Cohen’s-d (i.e., mean change/standard deviation of change, reporting bootstraped means and 95% CIs with 1000 resamples), comparing them to MDS-UPDRS scores. The MDS-UPDRS scores were those typically used as primary or secondary endpoints in PD clinical trials, including both ‘on-’ and ‘off-state’ MDS-UPDRS Parts I+II+III total, Parts II+III, and Part III.

Exploratory analyses also included determining the earliest time point for progression detection based on within-participant changes over time in the pooled PPP-development plus -test cohorts. For a given measure, we simulated 100 studies, each randomly sampling 100 participants without replacement (mimicking the samples for wearable data collection in prior PD clinical trials). We recorded the earliest time point where a significant and persistent (sustained onward, until study end) within-participant change could be detected in each study. The median of these earliest time points across the 100 simulated studies was reported as the earliest time point for progression detection.

We replicated the primary and secondary analyses in the PPMI-PD cohort as exploratory external evaluation.

#### Specificity to PD diagnosis

As part of external evaluation, we investigated whether the changes in the digital measures were specific to a PD diagnosis, by comparing PD and non-PD cohorts. We estimated the rate of within-participant changes for each measure in the PPMI-PD cohort (rate_PD), and in the PBHS-non-PD cohort (rate_nonPD) using separate linear mixed effects models (same method as the primary analysis in PPP-test cohort) and BH-adjusted p-values of the monthly increments were compared to alpha-level of 0.05. The comparison enabled categorizing the estimated measure changes as: disease-progression changes only, disease-progression plus non-disease specific changes, non-disease specific changes only, and no disease-progression changes (Suppl. Table 3).

#### Correlations with Clinical Measures and Group Separability

As exploratory analyses, we investigated the measures’ cross-sectional association with conventional clinical measures and their separability in different PD-severity groups (Suppl. Methods).

## Results

### Participants

Participant characteristics in the cohorts PPP-development (N=153), PPP-test (N=101), PPMI-PD (N=67) and PBHS non-PD (N=171) are summarized in Table 1 (Suppl. Fig 2).

**Table 1.** Demographic and clinical characteristics of cohorts of interest in this study.

|  | PPP |  |  | PPMI | PBHS |  |
| --- | --- | --- | --- | --- | --- | --- |
|  | Main<br>(N=517) | Developmental (PPP-development)<br>(N=153) | Internal Validation<br>(PPP-test)<br>(N=101) | Ext.Eval<br>(PPMI-PD)<br>(N=67) | Main<br>(N=2502) | Matched<br>(PBHS non-PD)<br>(N=171) |
| **Baseline demographics** |  |  |  |  |  |  |
| Mean age, yrs (SD) | 61.7 (9.1) | 61.9 (8.5) | 62.1 (9.0) | 69.2 (7.8) | 50 (17.2) | 68.2 (8.6) |
| Gender men, n (%) | 306 (59.2) | 93 (60.8) | 51 (50.5) | 45 (67.2) | 1127 (45.0) | 107 (62.3) |
| Watch worn left, n (%) | 474 (91.7) | 143 (93.5) | 91 (90.1) | NA | NA | NA |
| Watch worn most affected side, n (%) | 207 (40.0) | 71 (46.4) | 35 (34.7) | NA | NA | NA |
| Not employed in paid job, n (%) | 351 (67.9%) | 105 (68.6) | 62 (61.4) | NA | NA | NA |
| **At baseline visit** |  |  |  |  |  |  |
| Mean time since diagnosis, yrs (SD) | 2.7 (1.5) | 2.1 (1.1) | 2.2 (1.1) | 6.5 (2.3) | NA | NA |
| Mean MDS-UPDRS Part I (SD) | 14.5 (7.3) | 14.1 (6.4) | 14.6 (7.4) | 7.7 (3.4) | NA | NA |
| Mean MDS-UPDRS Part II (SD) | 8.4 (5.9) | 7.2 (4.5) | 7.2 (5.5) | 10.5 (5.5) | NA | NA |
| Mean MDS-UPDRS Part III (SD) |  |  |  |  |  |  |
| ON state | 28.5 (12.3) | 25.3 (9.1) | 26.5 (11.5) | 25.5 (10.9) | NA | NA |
| OFF state | 33.4 (12.8) | 29.6 (9.0) | 31.4 (12.4) | 33.9 (14.3) | NA | NA |
| Hoehn and Yahr Scale, n (%) |  |  |  |  | NA | NA |
| 0 | 0 | 0 | 0 | 1 (1.5) |  |  |
| 1 | 46 (9) | 10 (6.7) | 9 (8.9) | 10 (15.4) |  |  |
| 2 | 402 (78.7) | 136 (90.7) | 77 (76.2) | 47 (72.3) |  |  |
| 3 | 57 (11.2) | 4 (2.7) | 15 (14.9) | 7 (10.8) |  |  |
| 4 | 6 (1.2) | 0 | 0 | 0 |  |  |
| 5 | 0 | 0 | 0 | 0 |  |  |
| Mean PDQ-39 (SD) | 7.3 (6.8) | 6.4 (5.4) | 6.9 (7.0) | N/A | NA | NA |
| Mean Schwab and England ADL (SD) | 90.6 (8.9) | 92.7 (5.7) | 92.2 (8.1) | 85.4 (8.8) | NA | NA |
| Dyskinesia observed in clinic, n (%) | 41 (7.9) | 5 (3.3) | 6 (5.9) | 15 (22.4) | NA | NA |
| On symptomatic PD medication, n (%) | 474 (91.7) | 137 (89.5) | 94 (93.1) | 65 (97.0) | NA | NA |
| ADL= Activities of daily living; MDS-UPDRS = Movement disorder society-sponsored unified Parkinson's disease rating scale; NA = not applicable; PDQ-39 = Parkinson's disease questionnaire - 39; SD = Standard deviation |  |  |  |  |  |  |

Device daily wear times in PPP-test and PPMI cohorts were high (median 22.2 and 22.8 hours, respectively) and consistent throughout the study (Suppl. Fig 3).

### Measure selection

We analyzed 51 individual measures^34^ in the PPP-development cohort (N=153). From these, we selected 14 individual measures of walking and non-walking as input features for the development of CMs (Methods; Fig. 1).

**Figure 1.**
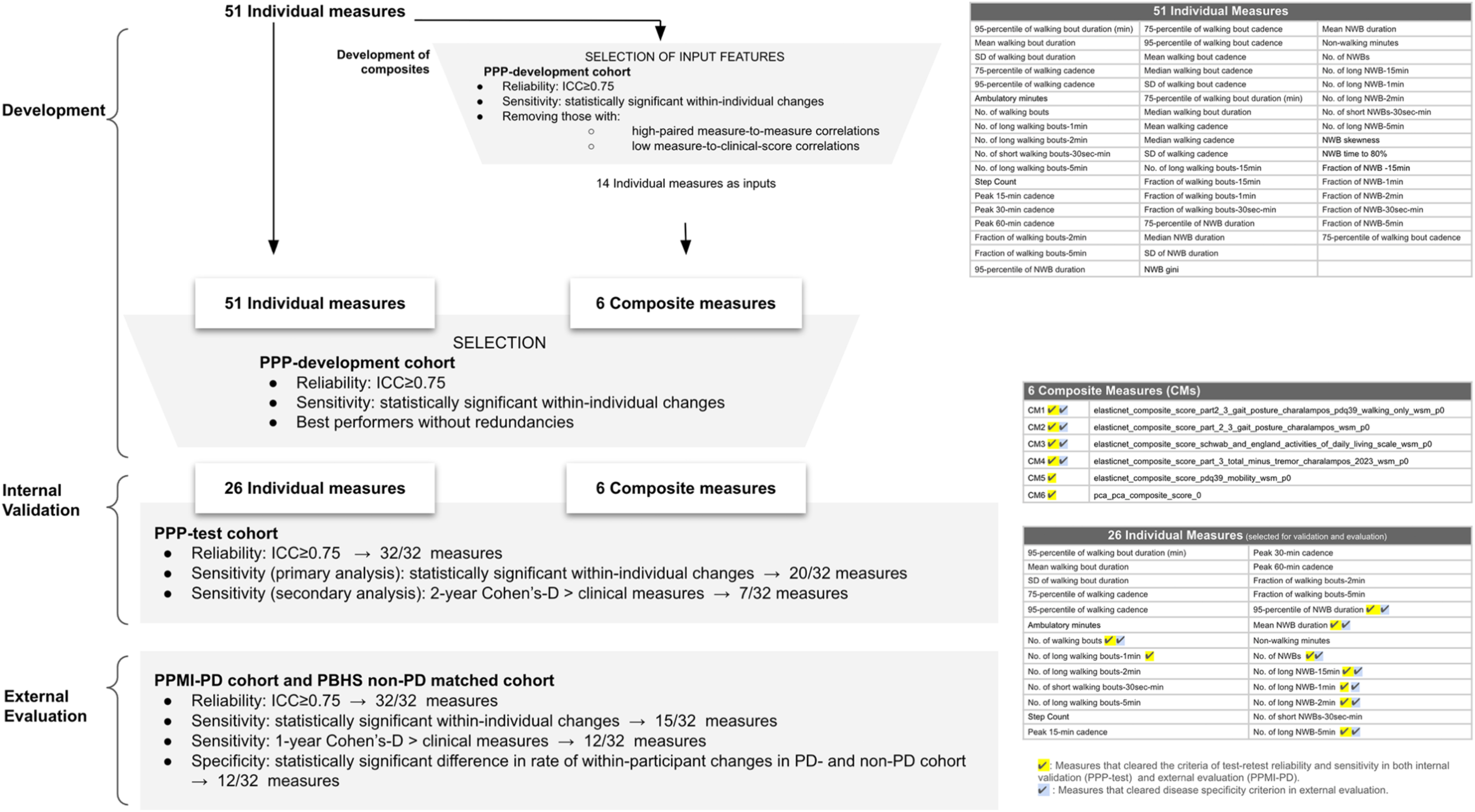
Overall schema for the selection, internal validation and external evaluation of measures.

To proceed to the internal validation and external evaluation on PPP-test and PPMI-PD datasets, we selected individual measures and CMs based on the criteria of test re-test reliability and sensitivity, removing those likely to be redundant. This process resulted in 32 candidate measures (26 individual, 6 composite; Fig. 1; Suppl. Table 1; Suppl. Data Tab 1).

### Measure sensitivity and quantification of disease progression

Full analysis results of the internal validation and external evaluation of the 32 measures can be found in Suppl. Data. This main section highlights a selection of 5 measures (3 individual and 2 composite) that we considered representative based on their domain (ie, walking and non-walking), the approach for their development (ie, individual, composite with supervised and unsupervised learning) and their performance results, for readability.

#### Internal validation

Twenty measures (14 individual, 6 composite) demonstrated significant within-individual changes in linear mixed models at the 1- and 2-year time points in the PPP-test cohort (N=101) (Suppl. Data Tab 2).

Seven digital measures showed larger effect size changes in this cohort than conventional clinic-based measures, which showed 2-year Cohen’s-d values ranging from 0.26 to 0.52 (Suppl. Fig. 5, Suppl. Data Tab 3). This was particularly pronounced for mean non-walking bout (NWB) duration (12.4% yearly change; 2-year Cohen’s-D, 0.623 [95% CI: 0.461,0.811]) and 95th percentile of NWB duration (17.1% yearly change; 2-year Cohen’s-D, 0.578 [95% CI: 0.420,0.782]), and the CM2 (2-year Cohen’s-D, 0.628 [95% CI: 0.411, 0.920]) (Fig. 2 A, B, Table 2).

**Figure 2.**
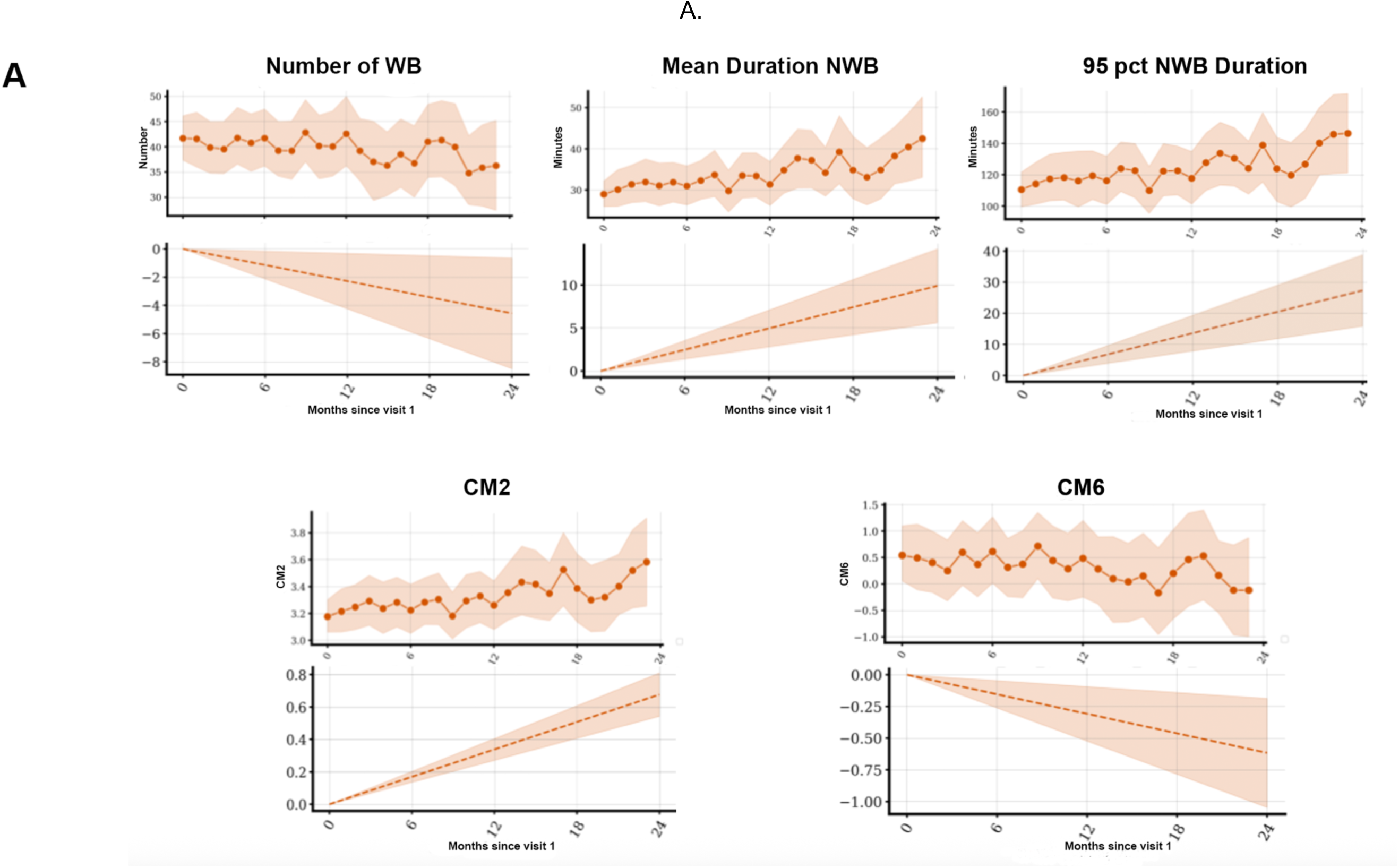
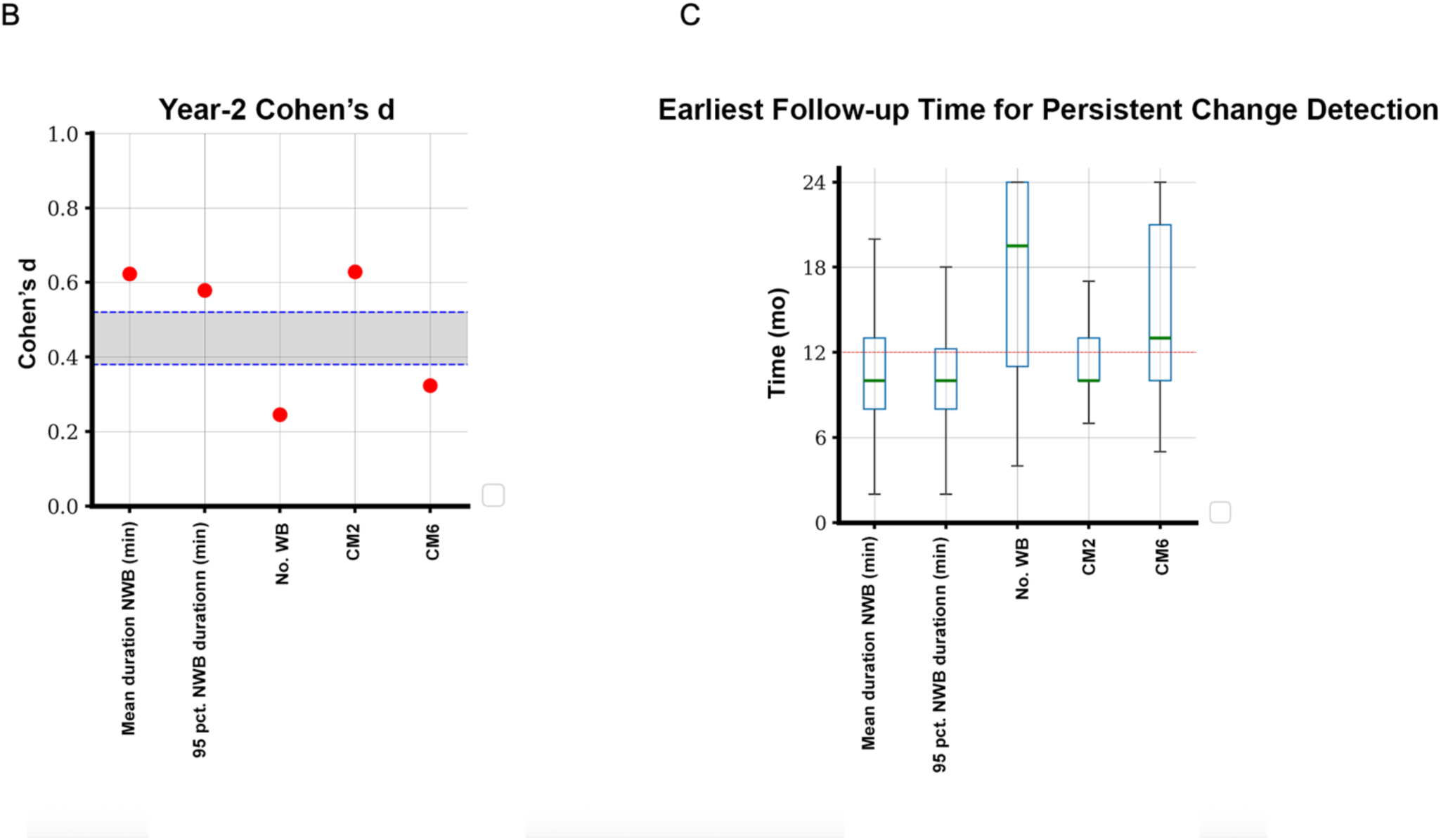
Internal validation results (PPP-test cohort based) of representative digital measures (see Suppl. Data Tabs 2-3, for full results). A. Monthly aggregated value in the PPP-test cohort over 2 years (top); mixed-effect model predicted rate of change over 2 years (bottom, shaded area: 95% CI of model predicted rate of change projected over time, see Supplementary Methods for note of CI derivation). B. Cohen’s-d analysis of changes from baseline at the 2-year follow-up point (shaded bracket: range of Cohen’s-d values at 2 years of select MDS-UPRS scores in the same participant cohort as reference). C. Median of earliest follow-up month at which significant and persistent measure change was detected (resampling 100 participants without replacement and repeated 100 times; horizontal dashed-line:12-month; boxes: 1st quartile (Q1) and 3rd quartile (Q3); error bars: Q1-1.5x inter-quartile range (IQR), Q3+1.5xIQR).

**Table 2.**
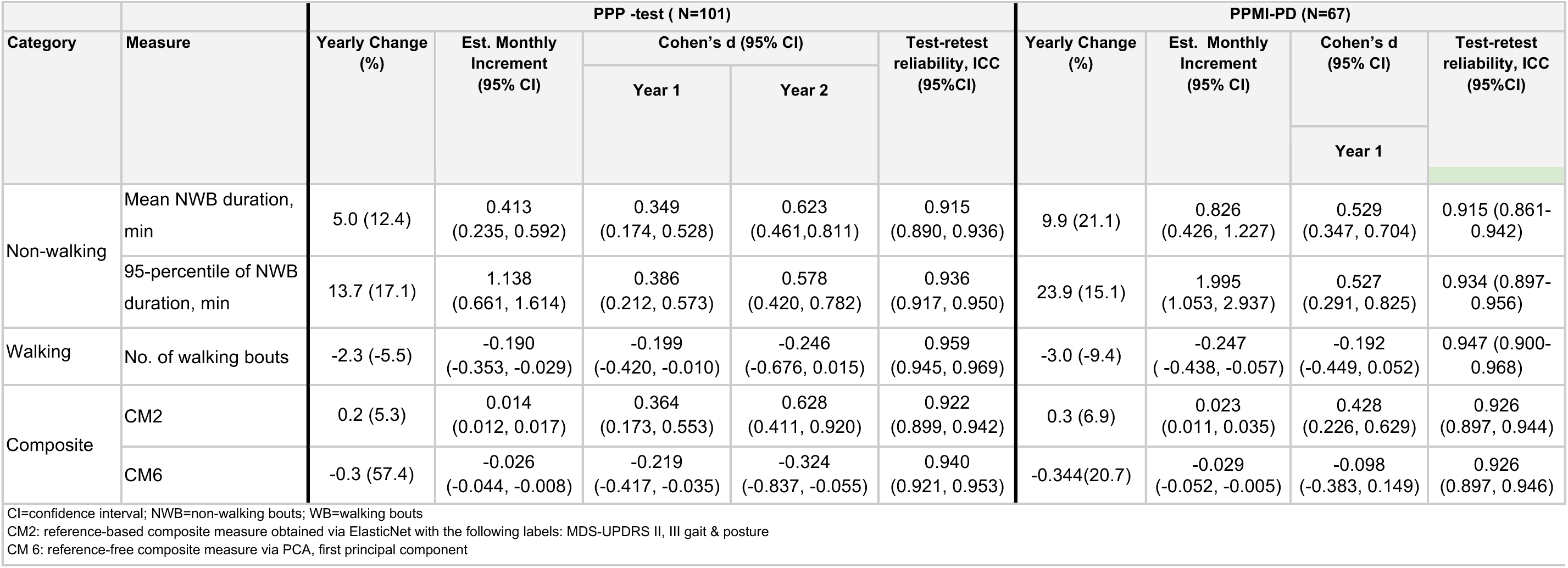
Summary of performance results of representative key digital measures in internal validation and external evaluation analyses (see Suppl. Data Tabs 2-5 for full results).

|  |  | PPP -test ( N=101) |  |  |  |  | PPMI-PD (N=67) |  |  |  |
| --- | --- | --- | --- | --- | --- | --- | --- | --- | --- | --- |
| Category | Measure | Yearly Change (%) | Est. Monthly Increment (95% CI) | Cohen's d (95% CI) |  | Test-retest reliability, ICC (95%CI) | Yearly Change (%) | Est. Monthly Increment (95% CI) | Cohen's d (95% CI) | Test-retest reliability, ICC (95%CI) |
|  |  |  |  | Year 1 | Year 2 |  |  |  | Year 1 |  |
| Non-walking | Mean NWB duration, min | 5.0 (12.4) | 0.413 (0.235, 0.592) | 0.349 (0.174, 0.528) | 0.623 (0.461,0.811) | 0.915 (0.890, 0.936) | 9.9 (21.1) | 0.826 (0.426, 1.227) | 0.529 (0.347, 0.704) | 0.915 (0.861-0.942) |
|  | 95-percentile of NWB duration, min | 13.7 (17.1) | 1.138 (0.661, 1.614) | 0.386 (0.212, 0.573) | 0.578 (0.420, 0.782) | 0.936 (0.917, 0.950) | 23.9 (15.1) | 1.995 (1.053, 2.937) | 0.527 (0.291, 0.825) | 0.934 (0.897-0.956) |
| Walking | No. of walking bouts | -2.3 (-5.5) | -0.190 (-0.353, -0.029) | -0.199 (-0.420, -0.010) | -0.246 (-0.676, 0.015) | 0.959 (0.945, 0.969) | -3.0 (-9.4) | -0.247 ( -0.438, -0.057) | -0.192 (-0.449, 0.052) | 0.947 (0.900-0.968) |
| Composite | CM2 | 0.2 (5.3) | 0.014 (0.012, 0.017) | 0.364 (0.173, 0.553) | 0.628 (0.411, 0.920) | 0.922 (0.899, 0.942) | 0.3 (6.9) | 0.023 (0.011, 0.035) | 0.428 (0.226, 0.629) | 0.926 (0.897, 0.944) |
|  | CM6 | -0.3 (57.4) | -0.026 (-0.044, -0.008) | -0.219 (-0.417, -0.035) | -0.324 (-0.837, -0.055) | 0.940 (0.921, 0.953) | -0.344(20.7) | -0.029 (-0.052, -0.005) | -0.098 (-0.383, 0.149) | 0.926 (0.897, 0.946) |
| CI=confidence interval; NWB=non-walking bouts; WB=walking bouts<br>CM2: reference-based composite measure obtained via ElasticNet with the following labels: MDS-UPDRS II, III gait & posture<br>CM 6: reference-free composite measure via PCA, first principal component |  |  |  |  |  |  |  |  |  |  |

For those same 20 measures with significant within-participant changes in linear mixed models, the median time point at which we first detected a significant (and subsequently persistent) effect was 10 months (range for mean duration NWB: 2-20 months; for 95th percentile of NWB duration: 2-18 months; Fig 2C, Suppl. Fig. 4).

An exploratory sensitivity analysis, adjusting for LEDD medication state, yielded consistent results with the main analysis (Suppl. Data Tab 2).

#### External evaluation and comparison with non-PD cohort

External evaluation in a PPMI-PD cohort (N=67) produced results largely consistent with the internal validation (Suppl. Data Tab 4; Fig. 3). Fifteen measures (9 individual, 6 composite, all of which had shown significant sensitivity in the PPP-test cohort; Fig.1) demonstrated significant within-individual changes in linear mixed models at the 1-year time point. Twelve measures showed effect sizes greater than conventional clinic-based measures (whose 1-year Cohen’s-d values ranged from 0.09 to 0.29) (Suppl. Fig. 5, Suppl. Data Tab 5).

**Figure 3.**
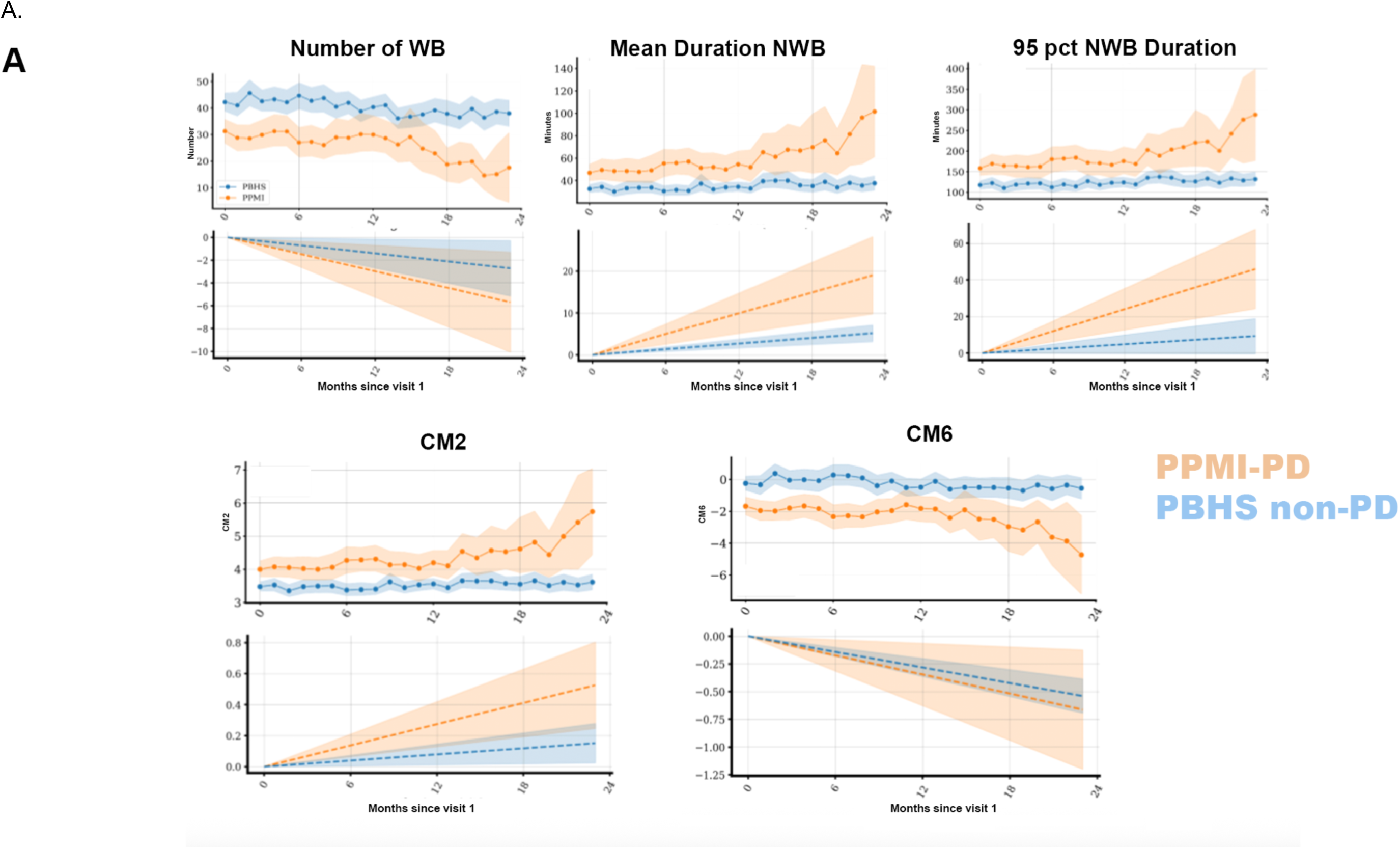
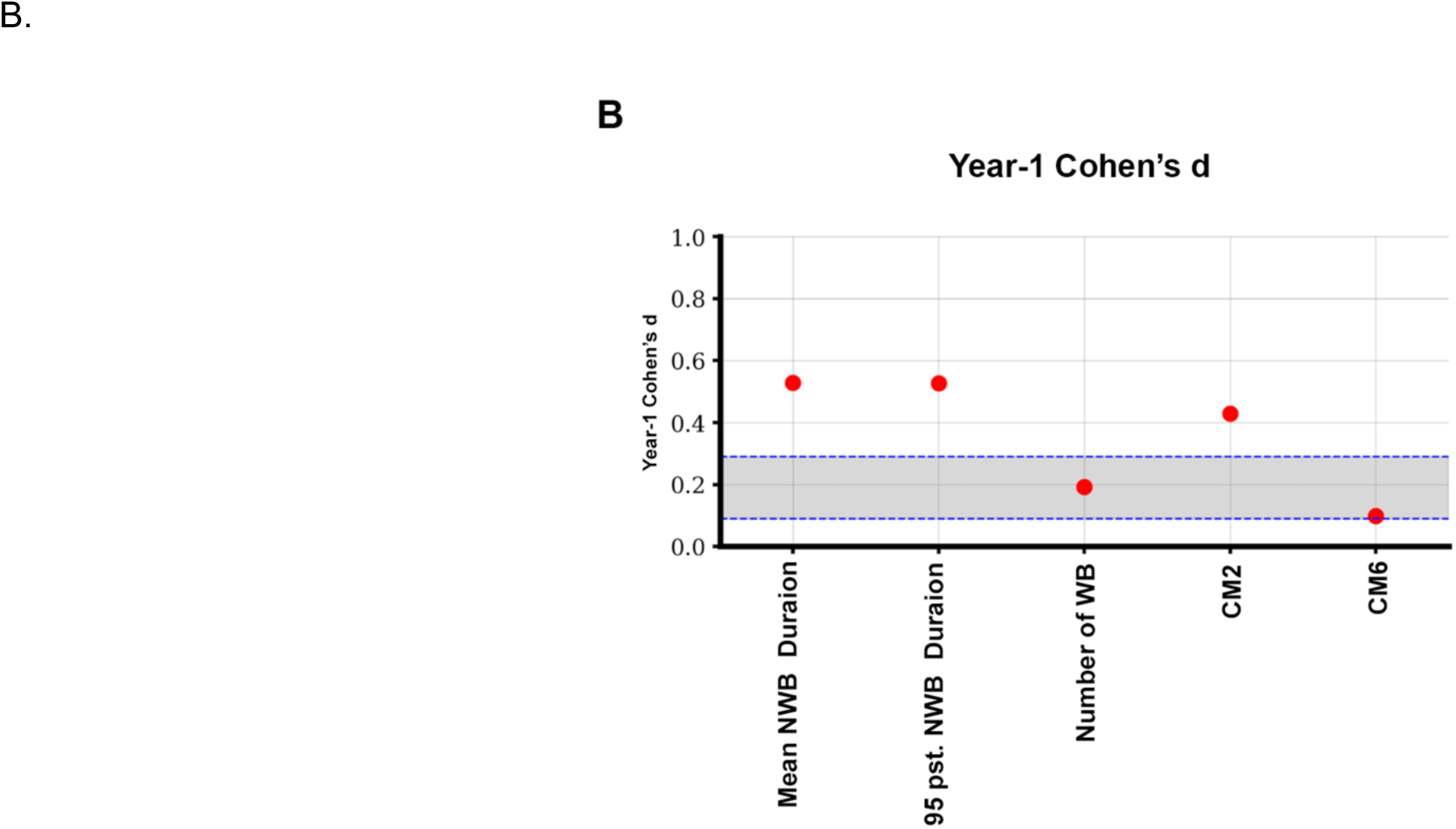
External evaluation results for select digital measures, PPMI-PD and PBHS non-PD cohorts (see Suppl. Data Tabs 4-5 for full results). A. Monthly aggregated value in the PPMI-PD and PBHS non-PD cohorts over 1 year (top); mixed-effect model predicted rate of change over 1 year (bottom, shaded area: 95% CI of model predicted rate of change projected over time). Note that, in this case, CM6 illustrates a measure that does not achieve a difference between the PPMI-PD and PBHS non-PD cohorts. B. Cohen’s-d analysis of changes at the 1-year follow-up point in the PD cohort (shaded bracket: range of Cohen’s-d values at 1 year of select MDS-UPRS scores in the same patient cohort as reference).

Similar to the PPP-test analysis, mean NWB duration (21.1% yearly change; 1-year Cohen’s-d, 0.529 [95%CI: 0.347, 0.704]) and 95th percentile NWB duration (15.1% yearly change; 1-year Cohen’s-d, 0.527 [95%CI: 0.291, 0.825]) showed the greatest longitudinal changes and Cohen-d values (Fig 3B, Table 2).

When comparing the PPMI-PD (N=67) and age/sex-matched non-PD (N=171) cohorts, 12 digital measures (8 individual, 4 composite) showed rates of longitudinal change greater in the PPMI-PD cohort than the non-PD cohort, directionally consistent with disease progression, thus differentiating disease progression trends from non-disease specific changes (Fig. 1; Suppl. Data Tab 4; Fig 3A).

##### Correlation with CROs and PROs

We observed weak-to-moderate correlations between the 32 digital measures and CROs/PROs in both the PPP-test (N=74) and PPMI-PD cohorts (N=58) (absolute-value Pearson r coefficients ranging from 0.002-0.48) (Suppl. Fig. 6). Composite measures appeared to have higher correlation with CROs/PROs than individual measures.

## Discussion

Wearable sensor-based digital measures could track changes in walking and non-walking behaviors in people with PD, over 12-24 months of daily living and with greater sensitivity than current gold-standard clinical measures. Digital measures detected significant and sustained changes from as early as 10 months of follow-up. Measures of non-walking behaviors (particularly NWB mean duration and 95th percentile of NWB duration) performed consistently well across analyses of test-retest reliability, sensitivity and specificity to PD, showing potential for use as disease progression markers in clinical trials.

Our study captures and quantifies sedentary behavior increases in people with early-stage PD, using continuous longitudinal data collected in real-life environments over years of disease progression. NWB duration-related measures showed the greatest sensitivity to PD progression, which aligns with clinical expectations and prior studies.^4–6^ The consolidation of sedentariness into longer non-walking bouts may happen as a functional consequence of multiple factors, such as postural instability and falling fears, or difficulties in initiating ambulation. It can also be a secondary manifestation of non-motor symptoms, such as depression, sensory or cognitive impairments.^43–45^

Our approach included CM development in addition to individual measures. Because CMs combine cross-domain signals into single metrics, they could be more comprehensive than individual measures.^46,47^ Our results suggest that CMs could have overall higher correlations with conventional CROs/PROs, which are meant to capture multi-domain aspects of PD symptoms. Both supervised and unsupervised learning approaches yielded reliable and sensitive CMs (according to our evaluation criteria), demonstrating the feasibility of developing CMs, with or without clinical scores as references. Although our findings are encouraging, additional research is warranted to establish a standard composite development framework, and potentially leverage other advanced, unsupervised machine learning techniques.

This study has unique strengths in the general context of research investigating digital measures in PD.^10,15–20,23–28,30,31^ Recent studies have evaluated passively collected digital measures’ sensitivity for PD progression^48^ and used exploratory digital endpoints in interventional trials.^33^ Ours is the first study with a comprehensive scope across meaningful aspects of patients’ health, by anchoring on multiple walking and non-walking behaviors reflective of overall mobility and physical function during daily living. This is also the first study to replicate results in independent datasets with relatively long follow-up and include a comparison with a non-PD cohort. The totality of our results confirms the sensitivity of our digital measures to detect disease-specific changes in a timely fashion. Our replication analyses also spanned various geographic regions, which enhances confidence about their potential generalizability.

We acknowledge limitations, and the need for future research. First, further contextualization of digital measures around known biological disease mechanisms is needed. For example, determining whether measures differ across the neuronal α-synuclein disease biological definition and integrated staging system (NSD-ISS) and/or groups with different genetic variants.^49,50^ Second, future research could delve into non-motor concepts, to develop a fuller picture of disease progression. Third, this study lacked essential anchors (e.g., patient-reported global impression of severity) to determine the minimal clinically important difference for digital measures, an aspect critical for future investigation. Finally, capturing data with a wearable device opens the question of whether the utility of these measures may be dependent on wear time or device adherence, a point that could be confirmed in future studies.

## Conclusions

Wearable sensor-based measures of walking and non-walking behaviors can quantify disease-specific changes during early-stage PD progression. The measures’ sensitivity, test re-test reliability, disease-specificity and reproducibility suggest potential application to develop clinical trial endpoints. The availability of measures such as these could facilitate clinical research by ushering in an era of smaller, less protracted and more patient-centric trials, based on sensitive metrics collected in patients’ own environments. This is particularly relevant in early-stage PD, where advances that accelerate the development of effective disease-modifying therapies could have the greater positive impact on patients’ lives.

## Supporting information

Supplementary methods and results

## Data Availability

The data supporting this study are not available for sharing. Requests for raw data from each of the originating studies can be directed to the sponsors.

## Disclosures/Acknowledgements page

### Study funding statement

This analysis was sponsored by Verily Life Sciences (Dallas, TX). Data used in the preparation of this article was obtained from the Personalized Parkinson Project (PPP), funded by Radboud University Medical Center, Radboud University, the city of Nijmegen and the Province of Gelderland, and the PPP Allowance made available by Health∼Holland, Top Sector Life Sciences & Health, to stimulate public-private partnerships.

Data used in the preparation of this article was obtained from the Project Baseline Health Study (PBHS), funded by Verily Life Sciences (Dallas, TX).

Data used in the preparation of this article was obtained on 2022-12-13 from the Parkinson’s Progression Markers Initiative (PPMI) database (www.ppmi-info.org/access-data-specimens/download-data), RRID:SCR_006431. For up-to-date information on the study, visit www.ppmi-info.org.

PPMI – a public-private partnership – is funded by the Michael J. Fox Foundation (MJFF) for Parkinson’s Research, and funding partners; including the Aligning Science Across Parkinson’s (ASAP) initiative. Other partners include 4D Pharma, Abbvie, AcureX, Allergan, Amathus Therapeutics, Aligning Science Across Parkinson’s, AskBio, Avid Radiopharmaceuticals, BIAL, BioArctic, Biogen, Biohaven, BioLegend, BlueRock Therapeutics, Bristol-Myers Squibb, Calico Labs, Capsida Biotherapeutics, Celgene, Cerevel Therapeutics, Coave Therapeutics, DaCapo Brainscience, Denali, Edmond J. Safra Foundation, Eli Lilly, Gain Therapeutics, GE HealthCare, Genentech, GSK, Golub Capital, Handl Therapeutics, Insitro, Jazz Pharmaceuticals, Johnson & Johnson Innovative Medicine, Lundbeck, Merck, Meso Scale Discovery, Mission Therapeutics, Neurocrine Biosciences, Neuron23, Neuropore, Pfizer, Piramal, Prevail Therapeutics, Roche, Sanofi, Servier, Sun Pharma Advanced Research Company, Takeda, Teva, UCB, Vanqua Bio, Verily, Voyager Therapeutics, the Weston Family Foundation and Yumanity Therapeutics.

### Study registration

Personalized Parkinson Project, clinicaltrials.gov ID NCT03364894; Parkinson’s Progression Markers Initiative, clinicaltrials.gov ID NCT04477785; Project Baseline Health Study, clinicaltrials.gov ID NCT03154346

### Prior disclosure of these data

Preliminary results related to these analyses were presented at the International Congress of Parkinson’s Disease and Movement Disorders; Philadelphia, PA; September 27, 2024.

### Authors’ contributions

Study concept and design: BRB, SS, RK, LJWE, KCH, SL

Data collection: Verily Life Sciences, PPP and PPMI; BRB, LHS, LCS, AS, LJWE

Data analysis and interpretation: KCH, SL, CSA, NK, ER, CC, WJM, RK, LJWE, BRB Draft writing and review: All

Draft approval for submission: All

## Acknowledgements

Authors wish to thank Hamsa Subramanian (formerly from Verily Life Sciences) for her valuable discussion and input to the non-PD cohort construction, Marjan J. Meinders (Radboud University) for her contributions to study conceptualization, Eric Gold (formerly from Verily Life Sciences) for project strategy and management support and Julia Saiz (Verily Life Sciences) for writing and editing support. Authors wish to thank participants, caregivers and study personnel for the Personalized Parkinson Project, the Parkinson’s Progression Markers Initiative and the Project Baseline Health Study.

