## Supplementary methods and results for "Wearable tracking of walking and non-walking as progression markers in early Parkinson’s disease"

#### Participant Selection Criteria

PPMI: Participants from the PPMI were eligible for this analysis without restrictions on disease duration, as most of them had been enrolled for more than 4 years when they entered into the Verily watch substudy.

PBHS: Excluded participants with own-medical or family history of musculoskeletal or neurological conditions.

#### Data Filtering

Inertial measurement unit (IMU) data preprocessing: Raw tri-axial accelerometer and gyroscope data was collected from Verily Study Watch embedded IMU. Standard signal preprocessing techniques are performed, including data resampling, signal bias removal, and noise removal using a band-pass filter. The preprocessed data was then used to derive digital measures.

### Data Selection, Availability and Processing

Days with analyzable data were defined as: those including in-bed periods for both nights, with between ≥8 and ≤24 hours of out-of-bed wear time, with daily number of bouts (see definition below) ≥2, where cadence is <3 steps/sec, with daily number of steps between ≥100 and ≤40000, and daily ambulatory minutes and non-walking daily minutes ≤24h. We filtered out days with >20 separate on-wrist segments (assuming this was likely due to a loose watch), where the on-wrist classification was performed by the Study Watch firmware on-wrist algorithm. Months with analyzable data were defined as: the first 24 months of wear time, months with ≥ 14 days with enough daily wear time, and from participants with at least 2 months of data.

### Composite Measure Development

Using the PPP-development dataset, individual measures were evaluated first against the clinical validation framework (Mixed-effect model and reliability tests) to identify a set of candidate individual measures (26 measures). Second, highly correlated individual measures were removed based on paired correlations ($\rho$>0.9) [30], resulting in the subset of individual measures used to build the composite measures (CMs). Third, two machine learning approaches were used to train CMs: supervised and unsupervised. Five supervised machine learning models were constructed and compared, which included lasso, ridge, Elastic Net, random forest, and gradient boosted regression tree. These models were trained to learn to aggregate the subset of individual measures into CMs. Five-fold cross validation was used to determine the optimal model hyperparameters. After evaluation, the elastic net model was selected as the primary modeling method, which utilized both L1 and L2 regularization to reduce overfitting. Five CMs were generated by training the elastic net model against five different labels: MDS-UPDRS II, III gait and PDQ-39 gait (CM1); MDS-UPDRS II, III gait & posture (CM2); Schwab & England ADL (CM3), MDS-UPDRS III (CM4), and PDQ-39 mobility (CM5). Clinical scores, such as MDS-UPDRS III could be subjective, or lack sensitivity or reliability [13], which could introduce bias to the supervised models. Therefore, we used an unsupervised learning approach, principal components analysis (PCA), that did not rely on clinical labels to guide the learning of the weights to aggregate the individual measures into composites, but identified independent principal components that were representative of the individual measures. Each principal component could be interpreted as a CM because a component captured various characteristics from multiple individual measures, without the guidance of any labels. The first principal component met the criteria of the clinical validation framework and it was used as CM6.

### Mixed Effect Model Specification to Estimate Sensitivity to Change

The null ($H_0$) and alternative ($H_A$) hypotheses of measure sensitivity can be stated as below:

$H_0$: rate_PD_$s$=0 vs $H_A$: rate_PD_$s$≠0, $s = 1, \dots, k$.

For simplicity of notation, the subscript $s = 1, \dots, k$ denoting the s$^{th}$ digital measure and it is dropped for the remainder of this document.

For a given digital measure, to test the above hypothesis, the raw monthly values of a digital measure are fitted using a linear mixed effects model. Below is the mathematical expression of the linear mixed effects model with raw monthly values of the digital measure taken as the response variable.

For participant $i = 1, \dots, N$ and month $j = 1, \dots, n_i$ ,

$$measure_{ij} = \gamma_{0i} + \gamma_{1i} time_{ij} + \gamma_{2ij} + e_{ij} \text{ where}$$

- $\gamma_{0i}$ consists of the time-invariant effects of age group, gender, baseline value of the given digital measure and the participant-specific random intercept $b_{0i}$, and is expressed as,

$$\gamma_{0i} = [\beta_{00} + b_{0i} + \beta_{01} I(gender_i = female) + \beta_{02} I(age\_grp_i = "65+") + \beta_{03} * I(YSD <= 2)]$$

- $\gamma_{1i}$ consists of the population and participant-specific monthly rate of progression, fixed slope $\beta_{10}$ and random slope $b_{1i}$, and is expressed as,

$\gamma_{1i} = [\beta_{10} + b_{1i}]$,

- $\gamma_{2ij}$ consist of the time-varying effects of seasonality, and COVID lockdown at a given time point j, where the reference level for seasonality is winter and for no lockdown for COVID-lockdown.

$$\gamma_{2ij} = [\beta_{20} I(season_{ij} = spring) + \beta_{21} I(season_{ij} = summer) + \beta_{22} I(season_{ij} = autumn)$$

$$+\beta_{23} I(lockdown_{ij} = mild) + \beta_{24} I(lockdown_{ij} = moderate) + \beta_{25} I(lockdown_{ij} = strict)$$

- and $e_i = (e_{ij})$ is a $n_i \times 1$ column vector of the residuals and assumed to be multivariate normal with zero mean and variance-covariance matrix $\sigma^2 I_{n_i}$,
- and $b_i = (b_{0i}, b_{1i})'$ is a $2 \times 1$ vector of the participant specific random intercept and random slope and assumed to multivariate normal with zero mean and variance-covariance matrix G, which is an unstructured variance-covariance matrix.

For a digital measure, if its model with random slope does not converge, a model with random intercept only will be rerun.

The measure sensitivity hypothesis testing for each digital measure can be restated in terms of $\beta_{10}$ as,

$$H_0: \beta_{10} = 0, vs\ H_A: \beta_{10} \neq 0.$$

The two-sided p-values resulting from the Wald test for $\beta_{10}$ from the linear mixed effects model is reported. To account for multiple testing of the measure sensitivity analysis in multiple digital measures, adjusted p-values are reported alongside the nominal p-value. The adjusted p-values are computed using the Benjamini-Hochberg (BH) procedure (Benjamini and Hochberg 1995) and the False Discovery Rate (FDR) will be set to 0.05. Statistical significance will be determined based on the BH adjusted p-values.

#### Mixed Effect Model Predicted Rate of Change CI derivation

The 95% CI displayed in Figures 1A and 1B (bottom panels) were derived as follows:

For mixed-effect model predicted rate of change over 2 years (i.e., $\hat{\beta} * t$), since 95% CI of $\hat{\beta} * t$ is $\hat{\beta}t \pm 1.96 * SE(\hat{\beta}t)$ and $SE(\hat{\beta}t) = Cov(\hat{\beta}t) = \sqrt{t^2\, Cov(\hat{\beta})} = t\sqrt{Cov(\hat{\beta})} = tSE(\hat{\beta})$, 95% CI of $\hat{\beta} * t$ becomes $\hat{\beta}t \pm 1.96 * t * SE(\hat{\beta})$. Thus with t ranging from 0 to 24 months, so the CI will become wider as time gets further away from time 0.

#### Levodopa Equivalent Daily Dose (LEDD)- Sensitivity Analysis

Because of the cyclic nature of the disease symptoms and medication effects, adjustments based on the assumption of any potential effect of LEDD on the digital measures were only considered as a sensitivity analysis to the primary analysis.

LEDD information was only collected at the three annual in-clinic visits, therefore this sensitivity analysis included only monthly aggregated digital measures computed from the three months before and after the in-clinic visits to ensure the temporal proximity and relevance of the LEDD status to the digital measures analyzed in the mixed effects model. This analysis was intended to stress test whether the results obtained in the primary analysis would be affected by the concurrent LEDD medication status. The LEDD analysis is performed by additionally including the time-varying LEDD (log2 transformed) in the primary analysis model specified previously.

### Correlation Analysis Between Digital Measures and Conventional Clinical Measures

As exploratory analysis, we investigated the measures' cross-sectional associations with conventional clinical measures collected during the baseline in-clinic visit (visit 1), separately for the PPP-test and PPMI-PD cohorts, using Pearson correlation coefficients. The conventional measures included CROs and PROs in the mobility and depression/apathy domains (Suppl. Table 4).

### Group Separability Analyses

We also conducted an exploratory analysis of the measures' separability in different PD-severity groups, defined by MDS-UPDRS scores and H&Y Scale, to test the hypothesis that they would show differences across relevant clinical groups. We used Kruskal Wallis tests to compare the group differences in digital measures, using only data points from baseline (clinical measures' first visit against digital measures' first evaluable month). Multiplicity control was not applied to this exploratory analysis. We performed analyses in eligible participants (those with evaluable baseline information) from a pooled PPP cohort (PPP-development plus PPP-test) and in the PPMI-PD cohort.

We tested the difference in the following groups based on conventional clinical measures: Hoehn & Yahr stages (H&Y), stage 1 vs 2 vs 3; off-state MDS-UPDRS part II+III scores, ≤40 vs >40 (LUMA, NCT05348785); and MDS-UPDRS part II total scores (from Neuronal Synuclein Disease Integrated Staging System [NDS-ISS][51]), ≤2 vs 3-13 vs >13 [49].

### Supplementary Results

#### Correlation with CROs and PROs

We observed weak-to-moderate correlations between the 32 digital measures and CROs/PROs in both the PPP-test and PPMI-PD cohorts (absolute values of Pearson r coefficients ranging from 0.002-0.48) (Suppl Fig 6). The highest correlations were between mean NWB duration or 95th percentile NWB duration and the Schwab&England ADL score in PPP-test.

#### Ability to distinguish clinically-relevant populations

We assessed whether digital measures showed separability across clinically-defined subgroups, in a pooled PPP-development plus -test cohort (N=213), and also in the PPMI-PD cohort (N=67). We found significant differences across clinical groups for most measures; for example, mean NWB duration ($P<0.001$) and 95th percentile NWB duration ($P=0.001$) separated in the MDS-UPDRS part II+III ≤40 vs >40 groups in the PPP-pooled cohort (Suppl. Fig 8).

**Supplementary Figure 1.** Design of the studies originating the cohorts relevant to our analyses.

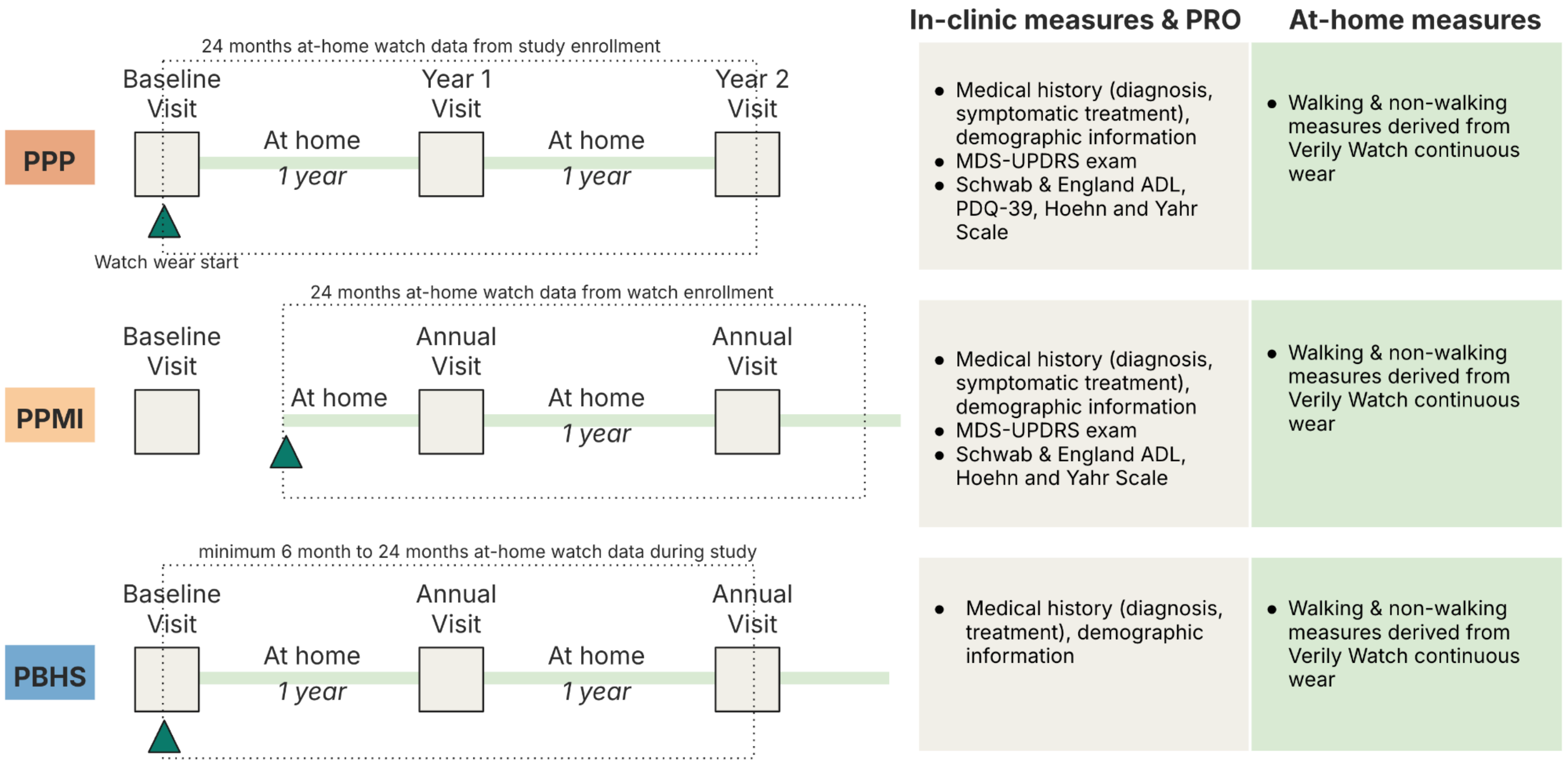

**Supplementary Figure 2.** Patient flow and selection for the cohorts of interest in the analyses.

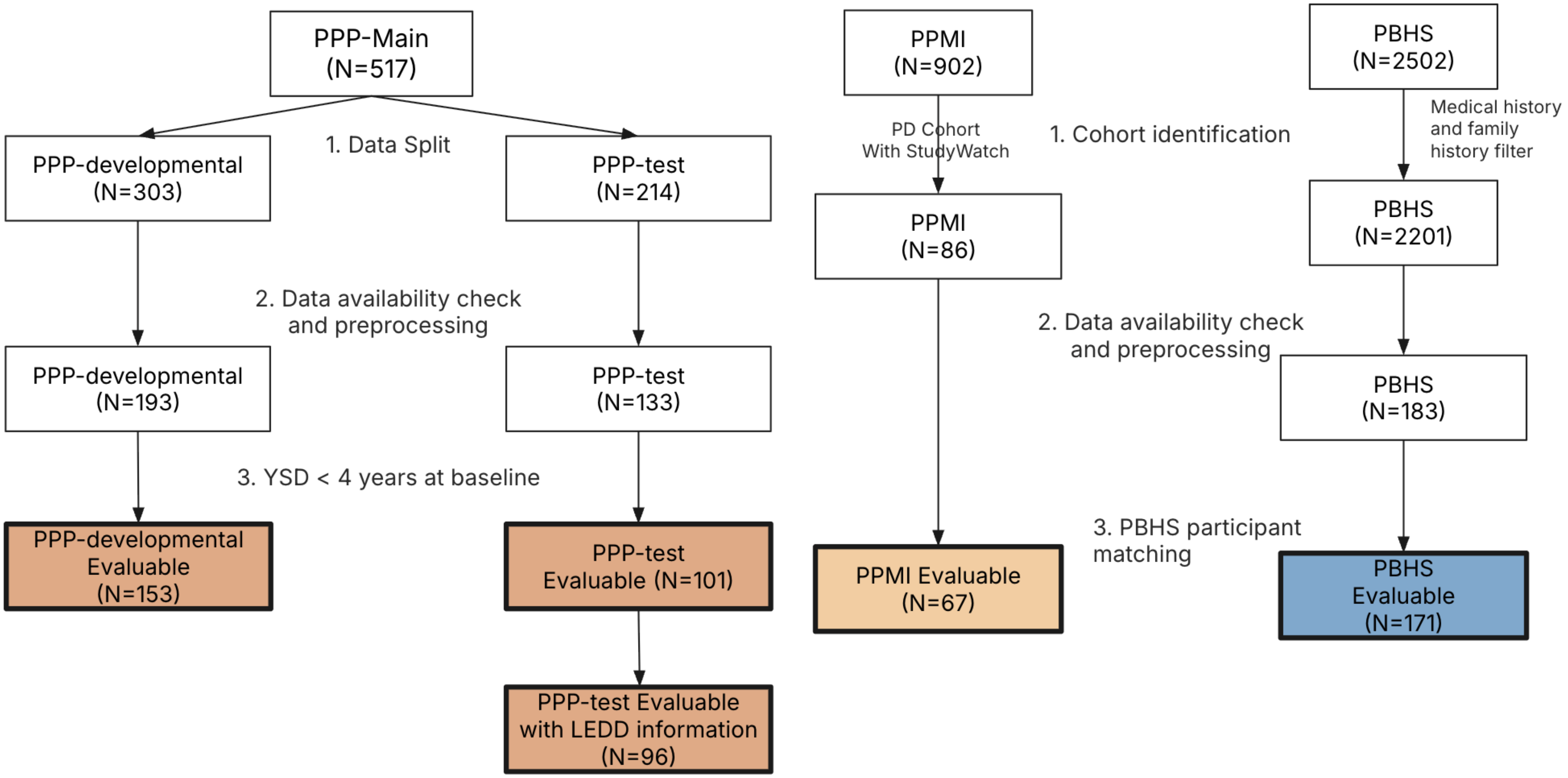

**Supplementary Figure 3.** Device wear time in the study cohorts.

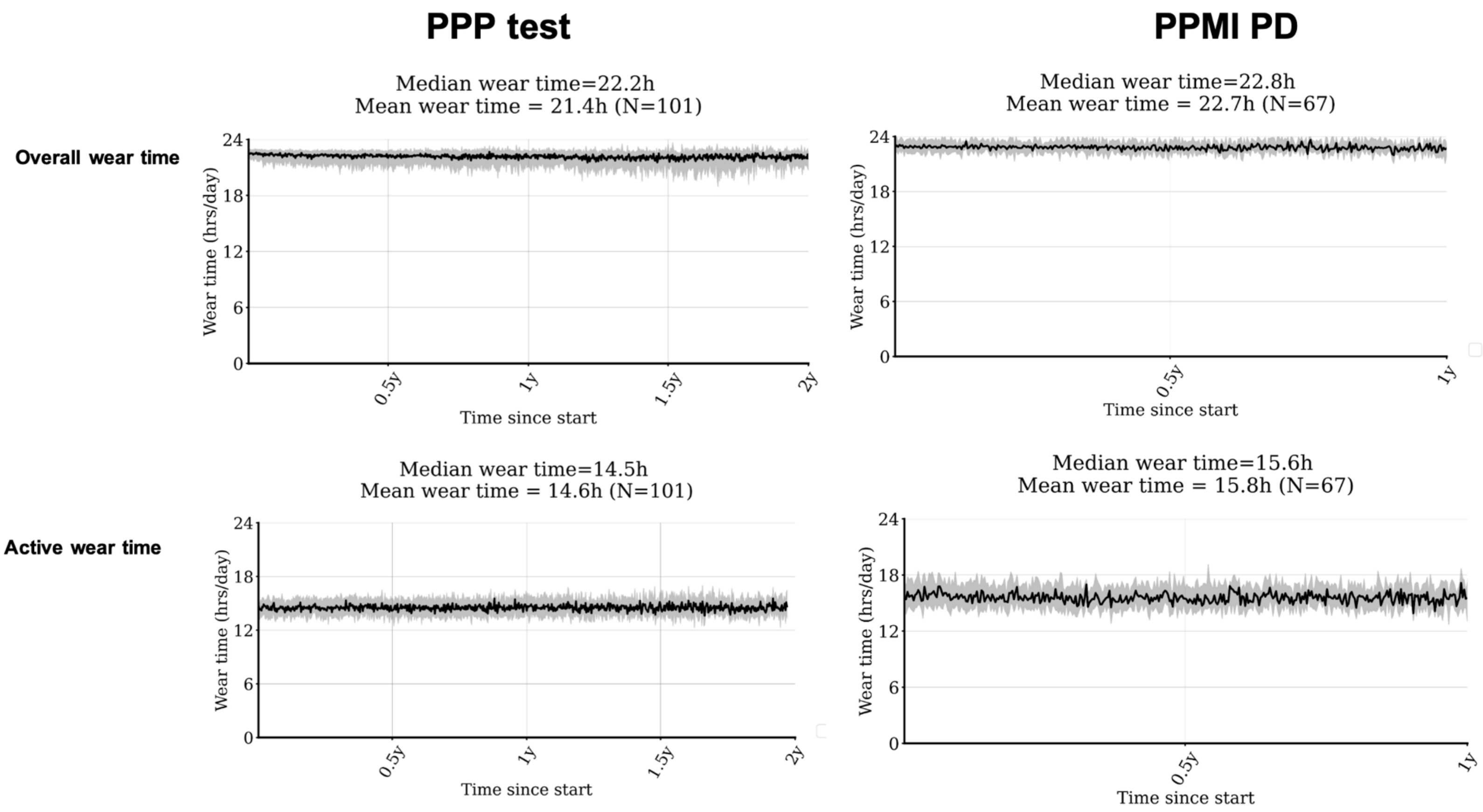

**Supplementary Figure 4.** Internal validation results in the PPP-test cohort for all investigated measures (see Suppl. Data for full results). A. Cohen's-d analysis of changes at the 1- and 2-year follow-up points (shaded bracket reflects range of Cohen's-d values of select MDS-UPRS scores in the same patient cohort). B. Earliest follow-up time point at which significant measure changes can be detected (horizontal dashed-line:12-month; boxes: 1st quartile (Q1) and 3rd quartile (Q3); error bars: Q1-1.5x inter-quartile range (IQR), Q3+1.5xIQR).

A.

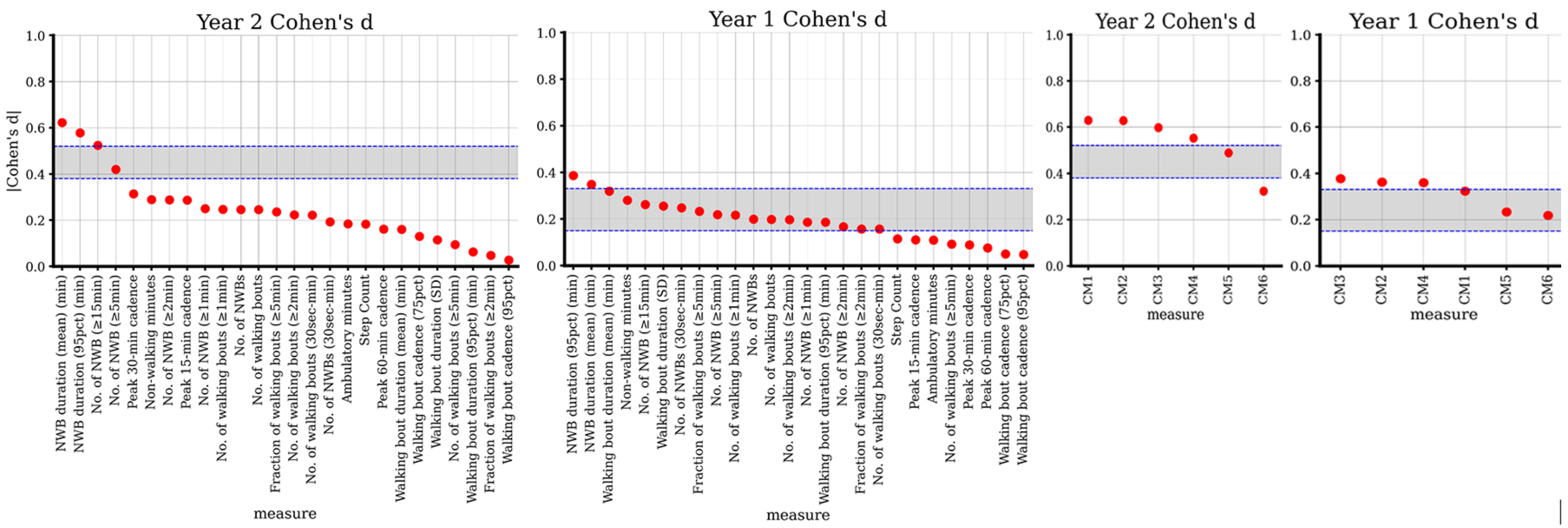

B.

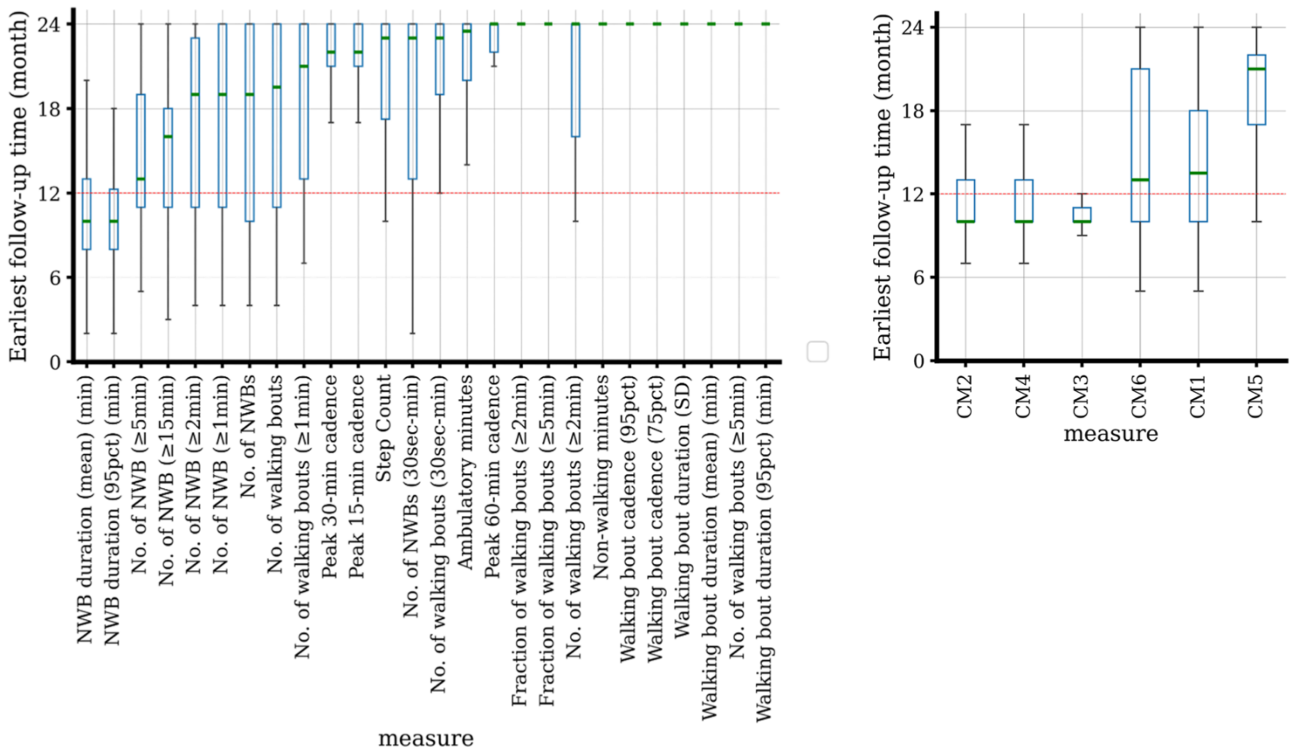
Earliest follow-up time (month)
24
18
12
6
0
NWB duration (mean) (min)
NWB duration (95pct) (min)
No. of NWB (≥5min)
No. of NWB (≥15min)
No. of NWB (≥2min)
No. of NWB (≥1min)
No. of NWBs
No. of walking bouts
No. of walking bouts (≥1min)
Peak 30-min cadence
Peak 15-min cadence
Step Count
No. of NWBs (30sec-min)
No. of walking bouts (30sec-min)
Ambulatory minutes
Peak 60-min cadence
Fraction of walking bouts (≥2min)
Fraction of walking bouts (≥5min)
No. of walking bouts (≥2min)
Non-walking minutes
Walking bout cadence (95pct)
Walking bout cadence (75pct)
Walking bout duration (SD)
Walking bout duration (mean) (min)
No. of walking bouts (≥5min)
Walking bout duration (95pct) (min)
measure
Earliest follow-up time (month)
24
18
12
6
0
CM2
CM4
CM3
CM6
CM1
CM5
measure

**Supplementary Figure 5.** External evaluation results, Cohen's-d analysis of 1-year follow-up point changes in the PPMI-PD cohort for all investigated digital measures (shaded bracket reflects range of Cohen's-D values at 1 year of select MDS-UPRS scores in the same patient cohort; see Suppl. Data for full results).

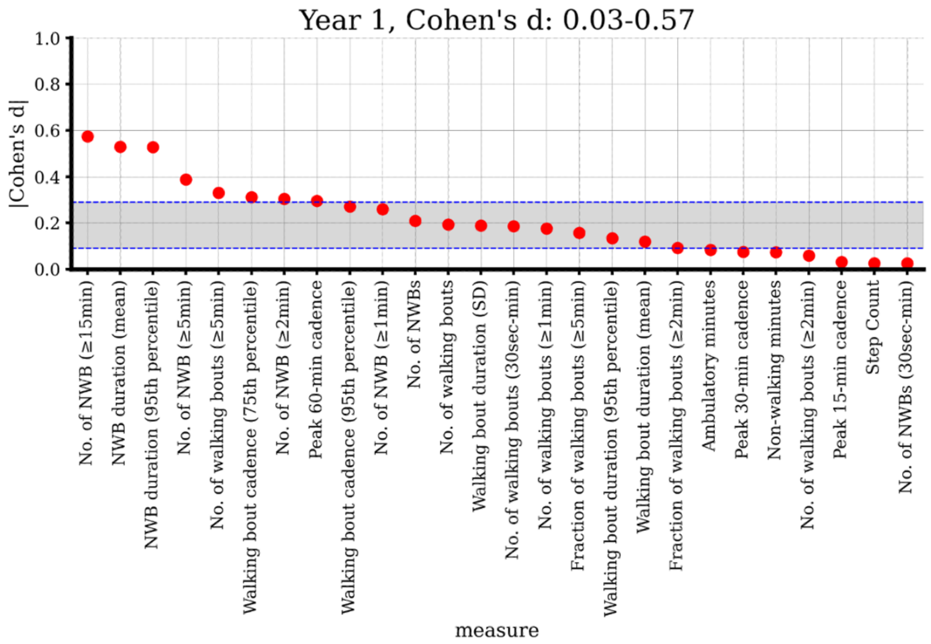


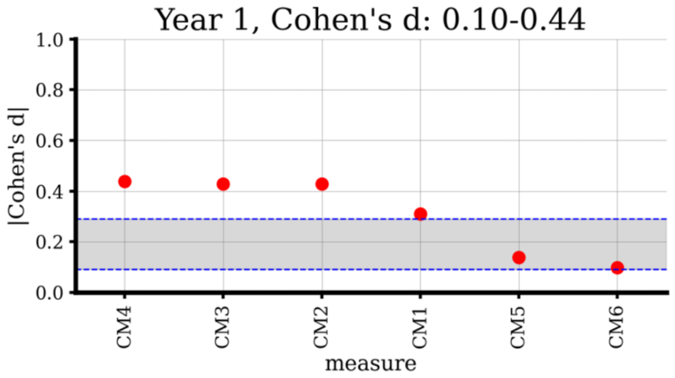

**Supplementary Figure 6.** Cross-sectional correlation analyses between digital measures and CROs and PROs. A. PPP-test cohort (N=74). B. PPMI-PD cohort (N=58)

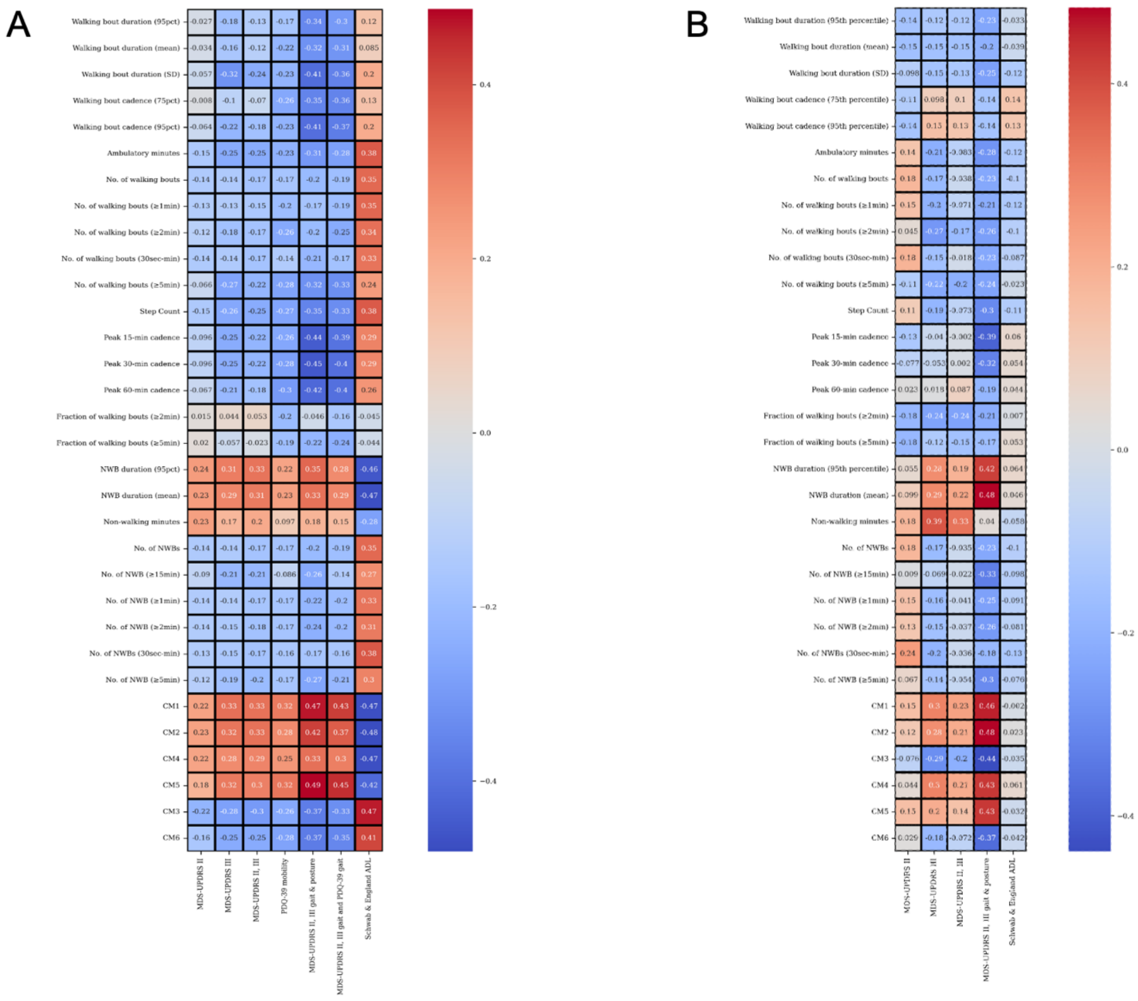

**Supplementary Figure 7.** Results from the group separability analysis on an example measure, mean NWB duration. A. Pooled PPP cohorts (-development and -test). B. PPMI-based cohort. Left: Groups based on the MDS-UPDRS Part II+III total (on-state) scores,≤40 and >40. Center: Groups based on MDS-UPDRS Part II scores, MDS-UPDRS Part II total ≤2, between ≥ 3 and ≤13, >13. Right: Hoehn & Yahr Stage 1, 2, 3

A. Pooled PPP cohorts (p<0.001, p=0.003, p=0.114, respectively)

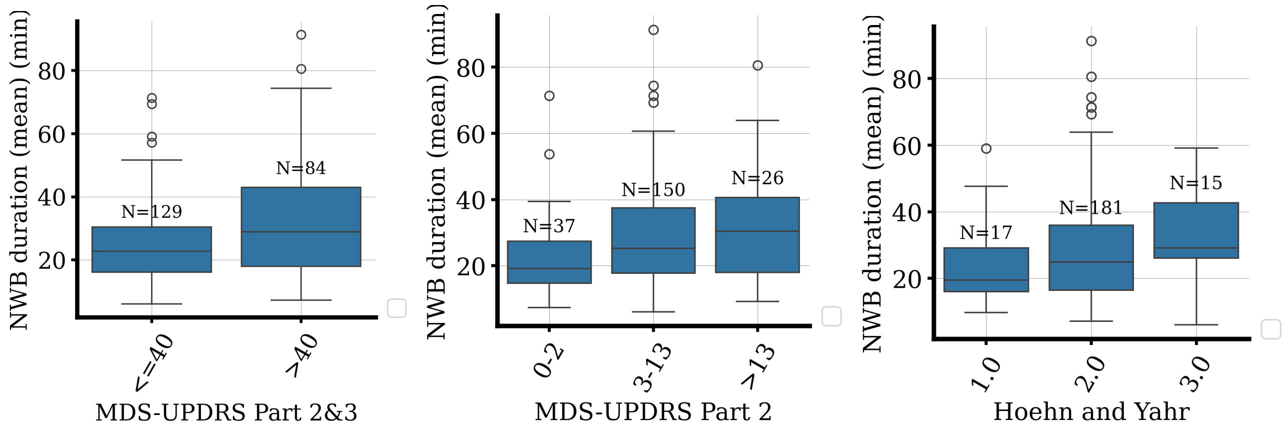

B. PPMI cohort (p=0.043, p=0.183, p=0.054, respectively)

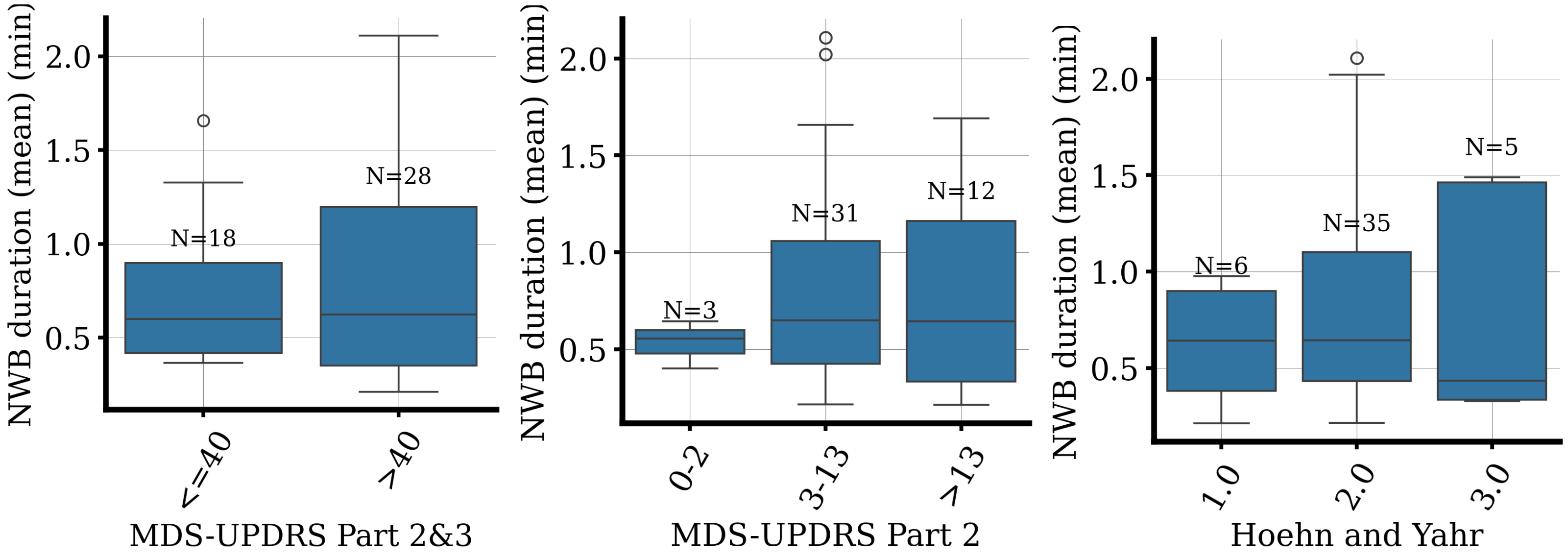

Supplementary Tables.

**Supplementary Table 1.** Definitions of the measures under investigation

| Measure components | Category | Type | Measure Name | Descriptions |
|---|---|---|---|---|
| 26 individual measures | Walking | Quantity | Step count | Summed number of steps per day |
| | | | Ambulatory minutes | Summed total minutes of walking per day |
| | | Bouts (WB) | Mean WB duration | Duration of daily walking bouts (≥30 seconds), mean value in minutes |
| | | | Standard deviation of WB duration | Duration of daily walking bouts (≥30 seconds), standard deviation in minutes |
| | | | 95-percentile of WB duration | Duration of daily walking bouts (≥30 seconds), 95 percentile value in minutes |
| | | | Number of WB | Number of daily walking bouts ( any duration ≥30 seconds) |
| | | | Number of short WB | Number of daily walking bouts (duration >30s to <1 minute) |
| | | | Number of long WB - 1 minute | Number of daily walking bouts (duration ≥1 minute) |
| | | | Number of long WB-2 minutes | Number of daily walking bouts (duration ≥2 minutes) |
| | | | Number of long WB-5 minutes | Number of daily walking bouts (duration ≥5 minutes) |
| | | | Fraction of long WB-2 minutes | Proportion of long bouts (≥2 minutes) out of total daily walking bouts |
| | | | Fraction of long WB-5 minutes | Proportion of long bouts (≥5 minutes) out of total daily walking bouts |
| | | Cadence | Peak cadence - 15 minutes | Average of the highest 10-sec cadence epochs of the day (for 15 minutes, not necessarily contiguous) |
| | | | Peak cadence - 30 minutes | Average of the highest 10-sec cadence epochs of the day (for 30 minutes, not necessarily contiguous) |

| Measure components | Category | Type | Measure Name | Descriptions |
|---|---|---|---|---|
| | | | Peak cadence - 60 minutes | Average of the highest 10-sec cadence epochs of the day (for 60 minutes, not necessarily contiguous) |
| | | | 75-percentile of cadence | Number of steps per second for the day, 75 percentile value |
| | | | 95-percentile of cadence | Number of steps per second for the day, 95 percentile value |
| | Non-walking | Quantity | Non-walking minutes | Summed total minutes of NWB (excluding sleep time) per day |
| | | Bouts (NWB) | Mean NWB duration | Duration of daily NWB (≥10 seconds), mean value in minutes |
| | | | 95-percentile of NWB duration | Duration of daily NWB (≥10 seconds), 95 percentile value in minutes |
| | | | Number of NWB | Number of NWB per day (any duration ≥10 seconds) |
| | | | Number of short NWB | Number of NWB per day (duration between 10s to <1 minute) |
| | | | Number of long NWB-1 minute | Number of NWB per day (duration ≥1 minutes) |
| | | | Number of long NWB-2 minutes | Number of NWB per day (duration ≥2 minutes) |
| | | | Number of long NWB-5 minutes | Number of NWB per day (duration ≥5 minutes) |
| | | | Number of long NWB-15 minutes | Number of NWB per day (duration ≥15 minutes) |
| 6 composite measures (CMs) | Reference-based composite measure | | CM1 | Elasticnet algorithm trained for MDS-UPDRS II, III gait and PDQ-39 gait) |
| | | | CM2 | Elasticnet algorithm trained for MDS-UPDRS II, III gait, posture |
| | | | CM3 | Elasticnet algorithm trained for Schwab & England ADL |
| | | | CM4 | Elasticnet algorithm trained for MDS-UPDRS III |
| | | | CM5 | Elasticnet algorithm trained for PDQ-39 mobility |
| | Reference-free composite measure | | CM6 | Product of the principal component analysis (PCA) with the largest variance (i.e., first principal component) |

**Supplementary Table 2.** Summary of the digital measure analyses performed in the study

<table>
<tr><th rowspan="2">Study stage</th><th rowspan="2">Cohort</th><th rowspan="2">Reliability, ICC</th><th colspan="3">Sensitivity, linear mixed-effect models</th><th colspan="2">Sensitivity, Cohen’s d</th><th rowspan="2">CRO/PRO correlation</th><th rowspan="2">Clinical group separability</th></tr>
<tr><th>Core modeling</th><th>Earliest time point with persisting significant change</th><th>Comparison vs non-PD</th><th>Core calculation</th><th>Comparison clinic-based measures</th></tr>
<tr><td>Development</td><td>PPP-development</td><td>✓</td><td>✓</td><td rowspan="2">✓<br>(pooled, exploratory)</td><td>-</td><td>✓</td><td>-</td><td>-</td><td rowspan="2">✓<br>(pooled, exploratory)</td></tr>
<tr><td>Internal validation</td><td>PPP-test</td><td>✓</td><td>✓<br>(primary)</td><td>-</td><td>✓<br>(secondary)</td><td>✓</td><td>✓<br>(exploratory)</td></tr>
<tr><td rowspan="2">External evaluation</td><td>PPMI-PD</td><td>✓</td><td>✓<br>(exploratory)</td><td>-</td><td rowspan="2">✓<br>(exploratory)</td><td>✓<br>(exploratory)</td><td>✓</td><td>✓<br>(exploratory)</td><td>✓<br>(exploratory)</td></tr>
<tr><td>PBHS non-PD</td><td>✓</td><td>✓<br>(exploratory)</td><td>-</td><td></td><td>-</td><td>-</td><td>-</td></tr>
</table>

**Supplement Table 3.** Rubric for the classification of estimated digital measure changes as disease-specific or not after the comparison between PPMI-PD and PBHS non-PD cohorts

<table>
<tr><th rowspan="3"></th><th colspan="4">Rate_nonPD</th></tr>
<tr><th colspan="2">Significant</th><th colspan="2">Non-significant</th></tr>
<tr><th>Inside of 95% CI</th><th>Outside of 95% CI</th><th>Inside of 95% CI</th><th>Outside of 95% CI</th></tr>
<tr><td>Rate_PD<br>Significant</td><td>Non-disease specific changes only</td><td>Disease progression plus non-specific changes</td><td>Non-disease specific changes only</td><td>Disease progression only</td></tr>
<tr><td>Non-significant</td><td colspan="4">No disease progression changes</td></tr>
</table>

Disease-progression changes only: rate_PD significant; rate_nonPD not significant; rate_PD outside 95% CI of rate_nonPD
Disease-progression plus non-disease specific changes: rate_PD and rate_nonPD both significant; rate_PD outside 95% CI of rate_nonPD
Non-disease specific changes only: rate_PD significant but rate_PD within 95% CI of rate_nonPD
No disease-progression changes: rate_PD not significant, regardless of rate_nonPD's significance

**Supplement Table 4.** Lists of conventional clinic-based CROs and PROs included in the correlation studies

| CROs | PROs |
|---|---|
| MDS-UPDRS II<br>MDS-UPDRS III<br>MDS-UPDRS II+III<br>MDS-UPDRS II+III gait and posture<br>Schwab & England ADL | PDQ-39 mobility<br>MDS-UPDRS II+III gait and PDQ-39 gait |
| MDS-UPDRS I | |
| MDS-UPDRS I, depression and anxiety moods, apathy | |

**Supplement Data Sheet (see separate file)**

Summary of performance results of all (32) digital measures across the different study stages: development (PPP-development cohort), internal validation (PPP-test cohort) and external evaluation (PPMI-PD and PBHS non-PD cohorts.
