## Supplementary Data for "Wearable tracking of walking and non-walking as progression markers in early Parkinson’s disease"

Key Performance Readouts for All Measures Investigated (51+6) in PPP-based Development Cohort (N=153)

| Category | Measure Name | 1-Year Cohen's d (95% CI) | 2-year Cohen's d (95% CI) | st. Monthly Increment (95% CI | FDR P-value | Test-retest ICC (95% CI) | Passed Criteria | SELECTED |
|---|---|---|---|---|---|---|---|---|
| Walking | 95-percentile of walking bout duration (min) | -0.006 (-0.136, 0.117) | 0.185 (0.043, 0.331) | 0.72 (0.198, 1.242) | 0.009 | 0.826 (0.771, 0.856) | Yes | Yes |
| Walking | Mean walking bout duration | -0.010 (-0.157, 0.109) | 0.268 (0.144, 0.394) | 0.252 (0.112, 0.392) | 0.001 | 0.832 (0.764, 0.866) | Yes | Yes |
| Walking | SD of walking bout duration | -0.040 (-0.165, 0.081) | 0.242 (0.110, 0.376) | 0.383 (0.129, 0.636) | 0.004 | 0.829 (0.773, 0.859) | Yes | Yes |
| Walking | 75-percentile of walking cadence | -0.147 (-0.259, -0.028) | -0.130 (-0.267, 0.014) | -0.001 (-0.001, 0.000) | 0.000 | 0.905 (0.883, 0.92) | Yes | Yes |
| Walking | 95-percentile of walking cadence | -0.145 (-0.273, -0.028) | -0.274 (-0.430, -0.132) | -0.001 (-0.001, -0.001) | 0.000 | 0.926 (0.904, 0.939) | Yes | Yes |
| Walking | Ambulatory minutes | -0.338 (-0.460, -0.220) | -0.495 (-0.686, -0.340) | -0.475 (-0.602, -0.348) | 0.000 | 0.89 (0.867, 0.912) | Yes | Yes |
| Walking | No. of walking bouts | -0.384 (-0.525, -0.254) | -0.675 (-0.867, -0.519) | -0.337 (-0.43, -0.243) | 0.000 | 0.943 (0.931, 0.952) | Yes | Yes |
| Walking | No. of long walking bouts-1min | -0.374 (-0.508, -0.256) | -0.673 (-0.852, -0.515) | -0.156 (-0.194, -0.118) | 0.000 | 0.899 (0.879, 0.915) | Yes | Yes |
| Walking | No. of long walking bouts-2min | -0.283 (-0.408, -0.155) | -0.567 (-0.762, -0.403) | -0.071 (-0.089, -0.054) | 0.000 | 0.843 (0.81, 0.87) | Yes | Yes |
| Walking | No. of short walking bouts-30sec-min | -0.322 (-0.460, -0.185) | -0.595 (-0.802, -0.439) | -0.178 (-0.250, -0.107) | 0.000 | 0.947 (0.934, 0.956) | Yes | Yes |
| Walking | No. of long walking bouts-5min | -0.177 (-0.295, -0.053) | -0.129 (-0.305, 0.019) | -0.013 (-0.016, -0.009) | 0.000 | 0.798 (0.747, 0.839) | Yes | Yes |
| Walking | Step Count | -0.337 (-0.459, -0.223) | -0.509 (-0.689, -0.360) | -50.393 (-64.973, -35.813) | 0.000 | 0.91 (0.89, 0.929) | Yes | Yes |
| Walking | Peak 15-min cadence | -0.323 (-0.430, -0.218) | -0.479 (-0.632, -0.348) | -0.002 (-0.003, -0.001) | 0.000 | 0.915 (0.894, 0.929) | Yes | Yes |
| Walking | Peak 30-min cadence | -0.306 (-0.407, -0.204) | -0.424 (-0.575, -0.295) | -0.002 (-0.003, -0.001) | 0.000 | 0.91 (0.889, 0.924) | Yes | Yes |
| Walking | Peak 60-min cadence | -0.208 (-0.313, -0.096) | -0.296 (-0.446, -0.163) | -0.002 (-0.002, -0.001) | 0.000 | 0.907 (0.885, 0.921) | Yes | Yes |
| Walking | Fraction of walking bouts-2min | -0.131 (-0.262, -0.006) | -0.023 (-0.173, 0.128) | 0.000 (0.000, 0.000) | 0.022 | 0.804 (0.767, 0.835) | Yes | Yes |
| Walking | Fraction of walking bouts-5min | -0.016 (-0.151, 0.104) | 0.271 (0.131, 0.421) | 0.000 (0.000, 0.000) | 0.000 | 0.792 (0.731, 0.829) | Yes | Yes |
| Non-walking | 95-percentile of NWB duration | 0.449 (0.332, 0.555) | 0.726 (0.606, 0.863) | 1.25 (0.902, 1.592) | 0.000 | 0.929 (0.907, 0.942) | Yes | Yes |
| Non-walking | Mean NWB duration | 0.467 (0.366, 0.566) | 0.645 (0.543, 0.759) | 0.415 (0.296, 0.535) | 0.000 | 0.919 (0.886, 0.937) | Yes | Yes |
| Non-walking | Non-walking minutes | 0.120 (0.003, 0.247) | 0.180 (0.035, 0.319) | 0.837 (0.29, 1.384) | 0.004 | 0.889 (0.866, 0.905) | Yes | Yes |
| Non-walking | No. of NWBs | -0.399 (-0.541, -0.268) | -0.691 (-0.887, -0.537) | -0.342 (-0.435, -0.249) | 0.000 | 0.943 (0.931, 0.952) | Yes | Yes |
| Non-walking | No. of long NWB-15min | -0.313 (-0.439, -0.197) | -0.774 (-0.950, -0.627) | -0.049 (-0.054, -0.043) | 0.000 | 0.913 (0.897, 0.925) | Yes | Yes |
| Non-walking | No. of long NWB-1min | -0.416 (-0.556, -0.284) | -0.701 (-0.911, -0.543) | -0.243 (-0.309, -0.177) | 0.000 | 0.946 (0.935, 0.955) | Yes | Yes |
| Non-walking | No. of long NWB-2min | -0.413 (-0.542, -0.283) | -0.734 (-0.937, -0.574) | -0.174 (-0.221, -0.127) | 0.000 | 0.945 (0.933, 0.954) | Yes | Yes |
| Non-walking | No. of short NWBs-30sec-min | -0.286 (-0.422, -0.164) | -0.571 (-0.715, -0.437) | -0.069 (-0.089, -0.048) | 0.000 | 0.904 (0.887, 0.918) | Yes | Yes |
| Non-walking | No. of long NWB-5min | -0.390 (-0.524, -0.261) | -0.745 (-0.960, -0.590) | -0.104 (-0.131, -0.076) | 0.000 | 0.936 (0.923, 0.946) | Yes | Yes |
| Composite | CM1 | 0.426 (0.322, 0.541) | 0.618 (0.503, 0.748) | 0.031 (0.027, 0.035) | 0.000 | 0.882 (0.842, 0.911) | Yes | Yes |
| Composite | CM2 | 0.490 (0.385, 0.604) | 0.697 (0.580, 0.832) | 0.018 (0.015, 0.02) | 0.000 | 0.898 (0.864, 0.924) | Yes | Yes |
| Composite | CM3 | -0.495 (-0.611, -0.383) | -0.754 (-0.897, -0.630) | -0.058 (-0.064, -0.051) | 0.000 | 0.922 (0.899, 0.938) | Yes | Yes |
| Composite | CM4 | 0.484 (0.366, 0.612) | 0.690 (0.571, 0.828) | 0.070 (0.063, 0.078) | 0.000 | 0.916 (0.893, 0.934) | Yes | Yes |
| Composite | CM5 | 0.364 (0.256, 0.474) | 0.529 (0.401, 0.674) | 0.037 (0.027, 0.047) | 0.000 | 0.893 (0.867, 0.913) | Yes | Yes |
| Composite | CM6 | -0.414 (-0.541, -0.295) | -0.705 (-0.889, -0.556) | -0.061 (-0.075, -0.046) | 0.000 | 0.902 (0.886, 0.918) | Yes | Yes |
| Walking | 75-percentile of walking bout cadence | -0.166 (-0.278, -0.055) | -0.146 (-0.288, -0.002) | -0.001 (-0.001, 0.000) | 0.000 | 0.86 (0.821, 0.883) | Yes | No |
| Walking | 95-percentile of walking bout cadence | -0.208 (-0.325, -0.098) | -0.254 (-0.396, -0.116) | -0.001 (-0.001, -0.001) | 0.000 | 0.887 (0.849, 0.906) | Yes | No |
| Walking | Mean walking bout cadence | -0.082 (-0.204, 0.040) | 0.031 (-0.108, 0.184) | 0.000 (0.000, 0.000) | 0.633 | 0.873 (0.843, 0.892) | No | No |
| Walking | Median walking bout cadence | -0.113 (-0.227, 0.011) | -0.058 (-0.199, 0.091) | 0.000 (-0.001, 0.000) | 0.088 | 0.861 (0.829, 0.881) | No | No |
| Walking | SD of walking bout cadence | -0.300 (-0.433, -0.173) | -0.585 (-0.776, -0.433) | -0.001 (-0.001, -0.001) | 0.000 | 0.875 (0.846, 0.895) | Yes | No |
| Walking | 75-percentile of walking bout duration (min) | -0.035 (-0.199, 0.093) | 0.171 (0.028, 0.289) | 0.236 (0.054, 0.418) | 0.014 | 0.768 (0.679, 0.812) | Yes | No |
| Walking | Median walking bout duration | -0.001 (-0.196, 0.104) | 0.166 (0.019, 0.266) | 0.066 (-0.007, 0.139) | 0.090 | 0.633 (0.496, 0.732) | No | No |
| Walking | Mean walking cadence | -0.055 (-0.172, 0.066) | -0.020 (-0.166, 0.136) | 0.000 (-0.001, 0.000) | 0.628 | 0.906 (0.885, 0.921) | No | No |
| Walking | Median walking cadence | -0.047 (-0.163, 0.076) | 0.081 (-0.063, 0.245) | 0.000 (0.000, 0.000) | 0.892 | 0.889 (0.867, 0.906) | No | No |
| Walking | SD of walking cadence | -0.233 (-0.354, -0.119) | -0.224 (-0.363, -0.077) | 0.000 (-0.001, 0.000) | 0.000 | 0.918 (0.897, 0.931) | Yes | No |
| Walking | No. of long walking bouts-15min | -0.033 (-0.151, 0.093) | 0.202 (0.061, 0.349) | 0.001 (-0.001, 0.003) | 0.337 | 0.765 (0.704, 0.809) | No | No |
| Walking | Fraction of walking bouts-15min | 0.088 (-0.041, 0.202) | 0.367 (0.250, 0.487) | 0.000 (0.000, 0.000) | 0.000 | 0.821 (0.705, 0.858) | Yes | No |
| Walking | Fraction of walking bouts-1min | -0.226 (-0.373, -0.103) | -0.219 (-0.397, -0.066) | -0.001 (-0.001, -0.001) | 0.000 | 0.793 (0.749, 0.822) | Yes | No |
| Walking | Fraction of walking bouts-30sec-min | 0.226 (0.103, 0.373) | 0.219 (0.066, 0.397) | 0.001 (0.001, 0.001) | 0.000 | 0.793 (0.749, 0.822) | Yes | No |
| Non-walking | 75-percentile of NWB duration | 0.413 (0.317, 0.509) | 0.575 (0.480, 0.686) | 0.570 (0.496, 0.645) | 0.000 | 0.895 (0.852, 0.918) | Yes | No |
| Non-walking | Median NWB duration | 0.340 (0.252, 0.422) | 0.480 (0.399, 0.562) | 0.240 (0.203, 0.277) | 0.000 | 0.845 (0.783, 0.875) | Yes | No |
| Non-walking | SD of NWB duration | 0.431 (0.319, 0.542) | 0.738 (0.614, 0.873) | 0.488 (0.351, 0.624) | 0.000 | 0.934 (0.914, 0.945) | Yes | No |
| Non-walking | NWB gini | 0.222 (0.100, 0.334) | 0.403 (0.245, 0.580) | 0.001 (0.001, 0.001) | 0.000 | 0.829 (0.802, 0.848) | Yes | No |
| Non-walking | NWB skewness | 0.080 (-0.045, 0.212) | -0.061 (-0.203, 0.094) | 0.000 (-0.001, 0.000) | 0.311 | 0.509 (0.412, 0.577) | No | No |
| Non-walking | NWB time to 80% | 0.365 (0.253, 0.480) | 0.692 (0.570, 0.825) | 1.307 (0.844, 1.770) | 0.000 | 0.92 (0.893, 0.935) | Yes | No |
| Non-walking | Fraction of NWB -15min | 0.403 (0.287, 0.520) | 0.572 (0.446, 0.712) | 0.002 (0.001, 0.003) | 0.000 | 0.905 (0.883, 0.92) | Yes | No |
| Non-walking | Fraction of NWB-1min | 0.229 (0.109, 0.347) | 0.288 (0.141, 0.448) | 0.001 (0.000, 0.001) | 0.000 | 0.84 (0.814, 0.861) | Yes | No |
| Non-walking | Fraction of NWB-2min | 0.281 (0.163, 0.398) | 0.398 (0.262, 0.547) | 0.001 (0.001, 0.001) | 0.000 | 0.875 (0.855, 0.892) | Yes | No |
| Non-walking | Fraction of NWB-30sec-min | -0.187 (-0.316, -0.057) | -0.345 (-0.496, -0.199) | -0.001 (-0.001, 0.000) | 0.000 | 0.813 (0.783, 0.837) | Yes | No |
| Non-walking | Fraction of NWB-5min | 0.341 (0.226, 0.453) | 0.509 (0.384, 0.648) | 0.002 (0.001, 0.002) | 0.000 | 0.898 (0.879, 0.912) | Yes | No |

Key Performance Readouts for All Measures Investigated (26+6) in PPP-based Test Cohort (N=101)

| Category | Measure Name | 1-Year Cohen's d (95% CI) | 2-year Cohen's d (95% CI) | st. Monthly Increment (95% CI | FDR P-value | Test-retest ICC (95% CI) | Passed Criteria based on PD cohort only | Est. Monthly Increment w. LEDD(95% CI) | FDR P-value w. LEDD | Passed Sensitivity Analysis (based on LEDD) |
|---|---|---|---|---|---|---|---|---|---|---|
| Walking | 95-percentile of walking bout duration (min) | 0.186 (-0.006, 0.363) | 0.064 (-0.219, 0.335) | 0.702 (-0.176, 1.579) | 0.138 | 0.842 (0.793, 0.88) | No | 1.283 (0.058, 2.507) | 0.055 | No |
| Walking | Mean walking bout duration (min) | 0.319 (0.169, 0.445) | 0.16 (-0.139, 0.397) | 0.365 (-0.041, 0.772) | 0.104 | 0.826 (0.786, 0.857) | No | 0.5 (0.023, 0.977) | 0.055 | No |
| Walking | SD of walking bout duration (min) | 0.256 (0.081, 0.405) | 0.115 (-0.133, 0.404) | 0.459 (-0.207, 1.124) | 0.191 | 0.85 (0.815, 0.878) | No | 0.576 (0.0, 1.152) | 0.065 | No |
| Walking | 75-percentile of walking cadence | -0.05 (-0.257, 0.141) | -0.131 (-0.435, 0.11) | 0.0 (-0.001, 0.001) | 0.354 | 0.914 (0.876, 0.937) | No | -0.001 (-0.001, 0.0) | 0.099 | No |
| Walking | 95-percentile of walking cadence | -0.048 (-0.24, 0.143) | -0.028 (-0.352, 0.227) | 0.0 (-0.001, 0.001) | 0.644 | 0.912 (0.863, 0.938) | No | 0.0 (-0.001, 0.0) | 0.265 | No |
| Walking | Ambulatory minutes | -0.11 (-0.316, 0.085) | -0.184 (-0.632, 0.09) | -0.257 (-0.539, 0.024) | 0.104 | 0.942 (0.924, 0.954) | No | -0.347 (-0.54, -0.154) | 0.001 | Yes |
| Walking | No. of walking bouts | -0.199 (-0.42, -0.01) | -0.246 (-0.676, 0.015) | -0.19 (-0.353, -0.027) | 0.042 | 0.959 (0.945, 0.969) | Yes | -0.209 (-0.311, -0.106) | 0.000 | Yes |
| Walking | No. of long walking bouts-1min | -0.216 (-0.383, -0.033) | -0.247 (-0.632, 0.008) | -0.096 (-0.166, -0.025) | 0.021 | 0.929 (0.902, 0.945) | Yes | -0.09 (-0.135, -0.044) | 0.000 | Yes |
| Walking | No. of long walking bouts-2min | -0.197 (-0.364, -0.011) | -0.223 (-0.583, 0.045) | -0.056 (-0.07, -0.042) | 0.000 | 0.861 (0.808, 0.894) | Yes | -0.036 (-0.056, -0.016) | 0.001 | Yes |
| Walking | No. of short walking bouts-30sec-min | -0.157 (-0.394, 0.033) | -0.222 (-0.625, 0.033) | -0.099 (-0.139, -0.058) | 0.000 | 0.961 (0.947, 0.971) | Yes | -0.117 (-0.183, -0.052) | 0.001 | Yes |
| Walking | No. of long walking bouts-5min | -0.093 (-0.29, 0.113) | -0.094 (-0.385, 0.168) | -0.008 (-0.016, 0.0) | 0.086 | 0.825 (0.759, 0.871) | No | -0.007 (-0.017, 0.002) | 0.146 | No |
| Walking | Step Count | -0.115 (-0.309, 0.08) | -0.184 (-0.615, 0.083) | -23.662 (-47.603, 0.279) | 0.086 | 0.929 (0.909, 0.945) | No | -33.32 (-50.617, -16.022) | 0.000 | Yes |
| Walking | Peak 15-min cadence | -0.111 (-0.317, 0.075) | -0.287 (-0.662, -0.012) | -0.001 (-0.002, 0.0) | 0.160 | 0.913 (0.871, 0.937) | No | -0.001 (-0.002, -0.0) | 0.003 | Yes |
| Walking | Peak 30-min cadence | -0.089 (-0.297, 0.099) | -0.314 (-0.713, -0.028) | -0.001 (-0.002, 0.0) | 0.131 | 0.914 (0.874, 0.937) | No | -0.001 (-0.002, -0.001) | 0.001 | Yes |
| Walking | Peak 60-min cadence | -0.076 (-0.27, 0.11) | -0.162 (-0.488, 0.111) | -0.001 (-0.002, -0.001) | 0.000 | 0.914 (0.876, 0.938) | Yes | -0.001 (-0.002, -0.0) | 0.008 | Yes |
| Walking | Fraction of walking bouts-2min | 0.158 (-0.033, 0.334) | -0.048 (-0.376, 0.201) | 0.0 (-0.001, 0.0) | 0.086 | 0.779 (0.726, 0.826) | No | 0.0 (-0.0, 0.001) | 0.535 | No |
| Walking | Fraction of walking bouts-5min | 0.233 (0.06, 0.395) | 0.236 (-0.039, 0.432) | 0.0 (0.0, 0.001) | 0.021 | 0.838 (0.788, 0.876) | Yes | 0.0 (-0.0, 0.001) | 0.087 | No |
| Non-walking | 95-percentile of NWB duration | 0.386 (0.212, 0.573) | 0.578 (0.42, 0.782) | 1.138 (0.661, 1.614) | 0.000 | 0.936 (0.917, 0.950) | Yes | 1.435 (1.094, 1.770) | 0.000 | Yes |
| Non-walking | Mean NWB duration | 0.349 (0.174, 0.528) | 0.623 (0.461, 0.811) | 0.413 (0.235, 0.592) | 0.000 | 0.915 (0.89, 0.936) | Yes | 0.510 (0.391, 0.629) | 0.000 | Yes |
| Non-walking | Non-walking minutes | 0.28 (0.109, 0.439) | 0.289 (0.038, 0.587) | 0.852 (0.236, 1.468) | 0.019 | 0.886 (0.862, 0.904) | Yes | 0.793 (0.331, 1.255) | 0.001 | Yes |
| Non-walking | No. of NWBs | -0.199 (-0.42, -0.012) | -0.246 (-0.68, 0.018) | -0.192 (-0.356, -0.029) | 0.042 | 0.959 (0.945, 0.969) | Yes | -0.384 (-1.074, 0.307) | 0.287 | Yes |
| Non-walking | No. of long NWB-15min | -0.262 (-0.486, -0.079) | -0.525 (-0.788, -0.299) | -0.032 (-0.052, -0.011) | 0.007 | 0.92 (0.899, 0.937) | Yes | -0.046 (-0.058, -0.033) | 0.000 | Yes |
| Non-walking | No. of long NWB-1min | -0.186 (-0.427, 0.005) | -0.25 (-0.674, 0.012) | -0.137 (-0.252, -0.023) | 0.040 | 0.959 (0.945, 0.968) | Yes | -0.148 (-0.222, -0.074) | 0.000 | Yes |
| Non-walking | No. of long NWB-2min | -0.167 (-0.402, 0.023) | -0.288 (-0.686, -0.037) | -0.099 (-0.131, -0.067) | 0.000 | 0.956 (0.943, 0.966) | Yes | -0.113 (-0.165, -0.061) | 0.000 | Yes |
| Non-walking | No. of short NWBs-30sec-min | -0.248 (-0.432, -0.07) | -0.193 (-0.571, 0.061) | -0.037 (-0.079, 0.004) | 0.104 | 0.94 (0.916, 0.955) | No | -0.038 (-0.063, -0.014) | 0.004 | Yes |
| Non-walking | No. of long NWB-5min | -0.218 (-0.468, -0.019) | -0.42 (-0.816, -0.164) | -0.061 (-0.079, -0.042) | 0.000 | 0.949 (0.935, 0.961) | Yes | -0.081 (-0.11, -0.053) | 0.000 | Yes |
| Composite | CM1 | 0.324 (0.148, 0.502) | 0.629 (0.372, 0.968) | 0.028 (0.023, 0.034) | 0.000 | 0.911 (0.882, 0.934) | Yes | -0.069 (-0.251, 0.113) | 0.509 | No |
| Composite | CM2 | 0.363 (0.173, 0.553) | 0.628 (0.411, 0.902) | 0.014 (0.012, 0.017) | 0.000 | 0.922 (0.899, 0.942) | Yes | 0.058 (0.044, 0.073) | 0.000 | Yes |
| Composite | CM3 | -0.378 (-0.578, -0.185) | -0.598 (-0.955, -0.355) | -0.049 (-0.07, -0.028) | 0.000 | 0.939 (0.921, 0.953) | Yes | 0.034 (0.022, 0.046) | 0.000 | Yes |
| Composite | CM4 | 0.36 (0.167, 0.556) | 0.552 (0.325, 0.897) | 0.049 (0.028, 0.071) | 0.000 | 0.938 (0.921, 0.952) | Yes | 0.028 (-0.074, 0.131) | 0.591 | No |
| Composite | CM5 | 0.234 (0.051, 0.418) | 0.489 (0.216, 0.868) | 0.029 (0.014, 0.044) | 0.000 | 0.912 (0.878, 0.935) | Yes | 0.016 (0.012, 0.02) | 0.000 | Yes |
| Composite | CM6 | -0.219 (-0.417, -0.035) | -0.324 (-0.837, -0.055) | -0.026 (-0.044, -0.008) | 0.005 | 0.94 (0.921, 0.953) | Yes | -0.031 (-0.043, -0.018) | 0.000 | Yes |

Key Performance Readouts for All Measures Investigated (26+6) in PPP-based Test Cohort (N=101)_Focus on the Secondary Analysis for Sensitivity (Cohen's-d)

| Category | Measure Name | 1-Year Cohen's d (95% CI) | Greater 1-y Cohen's d than best (highest Cohen's d) clinic-based measure? | 2-year Cohen's d (95% CI) | Greater 2-y Cohen's d than best (highest Cohen's d) clinic-based measures? |
|---|---|---|---|---|---|
| Walking | 95-percentile of walking bout duration (min) | 0.186 (-0.006, 0.363) | No | 0.064 (-0.219, 0.335) | No |
| Walking | Mean walking bout duration (min) | 0.319 (0.169, 0.445) | No | 0.16 (-0.139, 0.397) | No |
| Walking | SD of walking bout duration (min) | 0.256 (0.081, 0.405) | No | 0.115 (-0.133, 0.404) | No |
| Walking | 75-percentile of walking cadence | -0.05 (-0.257, 0.141) | No | -0.131 (-0.435, 0.11) | No |
| Walking | 95-percentile of walking cadence | -0.048 (-0.24, 0.143) | No | -0.028 (-0.352, 0.227) | No |
| Walking | Ambulatory minutes | -0.11 (-0.316, 0.085) | No | -0.184 (-0.632, 0.09) | No |
| Walking | No. of walking bouts | -0.199 (-0.42, -0.01) | No | -0.246 (-0.676, 0.015) | No |
| Walking | No. of long walking bouts-1min | -0.216 (-0.383, -0.033) | No | -0.247 (-0.632, 0.008) | No |
| Walking | No. of long walking bouts-2min | -0.197 (-0.364, -0.011) | No | -0.223 (-0.583, 0.045) | No |
| Walking | No. of short walking bouts-30sec-min | -0.157 (-0.394, 0.033) | No | -0.222 (-0.625, 0.033) | No |
| Walking | No. of long walking bouts-5min | -0.093 (-0.29, 0.113) | No | -0.094 (-0.385, 0.168) | No |
| Walking | Step Count | -0.115 (-0.309, 0.08) | No | -0.184 (-0.615, 0.083) | No |
| Walking | Peak 15-min cadence | -0.111 (-0.317, 0.075) | No | -0.287 (-0.662, -0.012) | No |
| Walking | Peak 30-min cadence | -0.089 (-0.297, 0.099) | No | -0.314 (-0.713, -0.028) | No |
| Walking | Peak 60-min cadence | -0.076 (-0.27, 0.11) | No | -0.162 (-0.488, 0.111) | No |
| Walking | Fraction of walking bouts-2min | 0.158 (-0.033, 0.334) | No | -0.048 (-0.376, 0.201) | No |
| Walking | Fraction of walking bouts-5min | 0.233 (0.06, 0.395) | No | 0.236 (-0.039, 0.432) | No |
| Non-walking | 95-percentile of NWB duration | 0.386 (0.212, 0.573) | Yes | 0.578 (0.42, 0.782) | Yes |
| Non-walking | Mean NWB duration | 0.349 (0.174, 0.528) | Yes | 0.623 (0.461, 0.811) | Yes |
| Non-walking | Non-walking minutes | 0.28 (0.109, 0.439) | No | 0.289 (0.038, 0.587) | No |
| Non-walking | No. of NWBs | -0.199 (-0.42, -0.012) | No | -0.246 (-0.68, 0.018) | No |
| Non-walking | No. of long NWB-15min | -0.262 (-0.486, -0.079) | No | -0.525 (-0.788, -0.299) | Yes |
| Non-walking | No. of long NWB-1min | -0.186 (-0.427, 0.005) | No | -0.25 (-0.674, 0.012) | No |
| Non-walking | No. of long NWB-2min | -0.167 (-0.402, 0.023) | No | -0.288 (-0.686, -0.037) | No |
| Non-walking | No. of short NWBs-30sec-min | -0.248 (-0.432, -0.07) | No | -0.193 (-0.571, 0.061) | No |
| Non-walking | No. of long NWB-5min | -0.218 (-0.468, -0.019) | No | -0.42 (-0.816, -0.164) | No |
| Composite | CM1 | 0.324 (0.148, 0.502) | No | 0.629 (0.372, 0.968) | Yes |
| Composite | CM2 | 0.363 (0.173, 0.553) | Yes | 0.628 (0.411, 0.902) | Yes |
| Composite | CM3 | -0.378 (-0.578, -0.185) | Yes | -0.598 (-0.955, -0.355) | Yes |
| Composite | CM4 | 0.36 (0.167, 0.556) | Yes | 0.552 (0.325, 0.897) | Yes |
| Composite | CM5 | 0.234 (0.051, 0.418) | No | 0.489 (0.216, 0.868) | No |
| Composite | CM6 | -0.219 (-0.417, -0.035) | No | -0.324 (-0.837, -0.055) | No |
| Clinic-based | MDS-UPDRS Part 1+2+3 total (Off-State) | 0.26 | | 0.5 | |
| Clinic-based | MDS-UPDRS Part 2+3 total (Off-State) | 0.32 | | 0.52 | |
| Clinic-based | MDS-UPDRS Part 3 total (Off-State) | 0.33 | | 0.48 | |
| Clinic-based | MDS-UPDRS Part 2 total | 0.15 | | 0.38 | |
| Clinic-based | MDS-UPDRS Part 1+2+3 total (On-State) | 0.22 | | 0.37 | |
| Clinic-based | MDS-UPDRS Part 2+3 total (On-State) | 0.27 | | 0.38 | |
| Clinic-based | MDS-UPDRS Part 3 total (On-State) | 0.21 | | 0.26 | |

Key Performance Readouts for All Measures Investigated (26+6) in PPMI-based PD Cohort (N=67) and PBHS-based non-PD cohort (N=171)

| | | PD Cohort (PPMI) | | | | | Non-PD cohort (PBHS) | | |
|---|---|---|---|---|---|---|---|---|---|
| Category | Measure Name | 1-Year Cohen's d (95% CI) | Est. Monthly Increment (95% CI) | FDR P-value | Test-retest ICC (95% CI) | Passed Criteria based on PD cohort only | Est. Monthly Increment (95% CI) | FDR P-value | Progression Trend Comparing PD cohort and Non-PD cohort |
| Walking | 95-percentile of walking bout duration (min) | 0.134 (-0.17, 0.342) | 1.187 (-1.213, 3.587) | 0.436 | 0.818 (0.636, 0.897) | No | 1.155 (0.435, 1.875) | 0.010 | No changes in PD |
| Walking | Mean walking bout duration | 0.12 (-0.181, 0.337) | 0.347 (-0.29, 0.984) | 0.394 | 0.816 (0.664, 0.893) | No | 0.147 (-0.118, 0.412) | 0.363 | No changes in PD |
| Walking | SD of walking bout duration | 0.188 (-0.097, 0.36) | 0.996 (-0.274, 2.266) | 0.216 | 0.799 (0.659, 0.879) | No | 0.338 (-0.4, 1.076) | 0.428 | No changes in PD |
| Walking | 75-percentile of walking cadence | 0.312 (0.092, 0.536) | 0.001 (0.000, 0.002) | 0.067 | 0.928 (0.897, 0.95) | No | -0.001 (-0.002, 0.000) | 0.067 | No changes in PD |
| Walking | 95-percentile of walking cadence | 0.271 (0.038, 0.481) | 0.000 (-0.002, 0.002) | 0.815 | 0.914 (0.88, 0.931) | No | -0.001 (-0.002, 0.000) | 0.007 | No changes in PD |
| Walking | Ambulatory minutes | -0.083 (-0.348, 0.154) | -0.3 (-0.605, 0.005) | 0.106 | 0.945 (0.903, 0.962) | No | -0.146 (-0.339, 0.048) | 0.201 | No changes in PD |
| Walking | No. of walking bouts | -0.192 (-0.449, 0.052) | -0.247 (-0.438, -0.057) | 0.029 | 0.947 (0.911, 0.963) | Yes | -0.118 (-0.223, -0.012) | 0.071 | disease progression (on top of non-disease specific changes (e.g., aging)) |
| Walking | No. of long walking bouts-1min | -0.175 (-0.528, 0.078) | -0.104 (-0.18, -0.028) | 0.023 | 0.889 (0.78, 0.937) | Yes | -0.078 (-0.124, -0.032) | 0.007 | Non-disease specific changes (e.g., aging) |
| Walking | No. of long walking bouts-2min | 0.059 (-0.253, 0.28) | -0.028 (-0.055, -0.001) | 0.095 | 0.832 (0.685, 0.888) | No | -0.032 (-0.052, -0.012) | 0.011 | No changes in PD |
| Walking | No. of short walking bouts-30sec-min | -0.186 (-0.438, 0.05) | -0.146 (-0.287, -0.005) | 0.091 | 0.957 (0.935, 0.968) | No | -0.066 (-0.1, -0.032) | 0.003 | No changes in PD |
| Walking | No. of long walking bouts-5min | 0.33 (0.13, 0.499) | 0.000 (-0.006, 0.007) | 0.928 | 0.783 (0.582, 0.847) | No | -0.004 (-0.011, 0.004) | 0.423 | No changes in PD |
| Walking | Step Count | 0.025 (-0.251, 0.25) | -17.19 (-43.753, 9.374) | 0.318 | 0.933 (0.885, 0.953) | No | -14.824 (-31.624, 1.977) | 0.127 | No changes in PD |
| Walking | Peak 15-min cadence | -0.031 (-0.336, 0.207) | -0.002 (-0.004, 0.001) | 0.259 | 0.915 (0.861, 0.942) | No | -0.001 (-0.002, 0.000) | 0.009 | No changes in PD |
| Walking | Peak 30-min cadence | 0.075 (-0.196, 0.327) | -0.001 (-0.003, 0.001) | 0.462 | 0.91 (0.868, 0.934) | No | -0.001 (-0.002, 0.000) | 0.034 | No changes in PD |
| Walking | Peak 60-min cadence | 0.295 (0.055, 0.534) | 0.000 (-0.001, 0.002) | 0.757 | 0.917 (0.882, 0.939) | No | -0.001 (-0.002, 0.000) | 0.105 | No changes in PD |
| Walking | Fraction of walking bouts-2min | 0.092 (-0.163, 0.369) | 0.000 (-0.001, 0.0) | 0.106 | 0.816 (0.666, 0.874) | No | 0.000 (-0.001, 0.000) | 0.206 | No changes in PD |
| Walking | Fraction of walking bouts-5min | 0.158 (-0.108, 0.382) | 0.000 (0.000, 0.000) | 0.564 | 0.821 (0.586, 0.898) | No | 0.000 (0.000, 0.000) | 0.020 | No changes in PD |
| Non-walking | 95-percentile of NWB duration | 0.527 (0.291, 0.825) | 1.995 (1.053, 2.937) | 0.001 | 0.934 (0.897, 0.956) | Yes | 0.402 (-0.016, 0.820) | 0.101 | Disease progression only |
| Non-walking | Mean NWB duration | 0.529 (0.347, 0.704) | 0.826 (0.426, 1.227) | 0.001 | 0.915 (0.861, 0.942) | Yes | 0.224 (0.138, 0.309) | 0.000 | disease progression (on top of non-disease specific changes (e.g., aging)) |
| Non-walking | Non-walking minutes | 0.073 (-0.182, 0.339) | -0.614 (-1.695, 0.466) | 0.391 | 0.934 (0.898, 0.949) | No | -0.316 (-0.785, 0.154) | 0.257 | No changes in PD |
| Non-walking | No. of NWBs | -0.208 (-0.468, 0.037) | -0.256 (-0.445, -0.066) | 0.024 | 0.947 (0.91, 0.963) | Yes | -0.115 (-0.22, -0.009) | 0.072 | Disease progression only |
| Non-walking | No. of long NWB-15min | -0.574 (-0.868, -0.331) | -0.076 (-0.113, -0.038) | 0.001 | 0.947 (0.918, 0.961) | Yes | -0.025 (-0.042, -0.007) | 0.025 | disease progression (on top of non-disease specific changes (e.g., aging)) |
| Non-walking | No. of long NWB-1min | -0.259 (-0.512, -0.018) | -0.216 (-0.362, -0.07) | 0.013 | 0.95 (0.921, 0.964) | Yes | -0.079 (-0.158, 0.000) | 0.093 | Disease progression only |
| Non-walking | No. of long NWB-2min | -0.304 (-0.552, -0.064) | -0.187 (-0.297, -0.077) | 0.004 | 0.953 (0.927, 0.965) | Yes | -0.06 (-0.12, 0.001) | 0.098 | Disease progression only |
| Non-walking | No. of short NWBs-30sec-min | -0.025 (-0.341, 0.225) | -0.028 (-0.062, 0.006) | 0.187 | 0.905 (0.814, 0.941) | No | -0.026 (-0.048, -0.005) | 0.055 | No changes in PD |
| Non-walking | No. of long NWB-5min | -0.387 (-0.649, -0.135) | -0.146 (-0.217, -0.076) | 0.001 | 0.953 (0.93, 0.964) | Yes | -0.039 (-0.079, 0.000) | 0.093 | Disease progression only |
| Composite | CM1 | 0.31 (0.09, 0.522) | 0.033 (0.01, 0.056) | 0.006 | 0.925 (0.898, 0.941) | Yes | 0.012 (0.001, 0.022) | 0.030 | disease progression (on top of non-disease specific changes (e.g., aging)) |
| Composite | CM2 | 0.428 (0.226, 0.629) | 0.023 (0.011, 0.035) | 0.000 | 0.926 (0.897, 0.944) | Yes | 0.007 (0.001, 0.012) | 0.029 | disease progression (on top of non-disease specific changes (e.g., aging)) |
| Composite | CM3 | -0.428 (-0.666, -0.21) | -0.075 (-0.113, -0.037) | 0.000 | 0.93 (0.907, 0.946) | Yes | -0.023 (-0.042, -0.004) | 0.031 | disease progression (on top of non-disease specific changes (e.g., aging)) |
| Composite | CM4 | 0.438 (0.218, 0.676) | 0.077 (0.038, 0.115) | 0.001 | 0.93 (0.905, 0.947) | Yes | 0.023 (0.004, 0.042) | 0.024 | disease progression (on top of non-disease specific changes (e.g., aging)) |
| Composite | CM5 | 0.138 (-0.102, 0.377) | 0.027 (0.009, 0.045) | 0.004 | 0.924 (0.897, 0.941) | Yes | 0.019 (0.004, 0.033) | 0.033 | Non-disease specific changes (e.g., aging) |
| Composite | CM6 | -0.098 (-0.383, 0.149) | -0.029 (-0.052, -0.005) | 0.016 | 0.926 (0.897, 0.946) | Yes | -0.023 (-0.030, -0.017) | 0.000 | Non-disease specific changes (e.g., aging) |

| Category | Definition |
|---|---|
| Disease progression only | PD-cohort shows statistically signficant change that meets clinical expectation, while non-PD cohort shows no change. |
| Disease progression (on top of non-disease specific changes (e.g., aging)) | PD and non-PD cohorts both shows significant change, but PD cohort shows statistically significant greater change. |
| Non-disease specific changes (e.g., aging) | PD and non-PD cohorts both shows significant change, but no difference between the no cohorts. |
| No changes in PD | PD cohort doesn't show significant change, regardless of non-PD cohort. |

Key Performance Readouts for All Measures Investigated (26+6) in PPMI-based PD Cohort (N=67) and PBHS-based non-PD cohort (N=171)_Focus on the Exploratory Analysis of Sensitivity (Cohen's-d)

| | | PD Cohort (PPMI) | |
|---|---|---|---|
| **Category** | **Measure Name** | **1-Year Cohen's d (95% CI)** | **Greater 1-y Cohen's d than clinic-based?** |
| Walking | 95-percentile of walking bout duration (min) | 0.134 (-0.17, 0.342) | No |
| Walking | Mean walking bout duration | 0.12 (-0.181, 0.337) | No |
| Walking | SD of walking bout duration | 0.188 (-0.097, 0.36) | No |
| Walking | 75-percentile of walking cadence | 0.312 (0.092, 0.536) | Yes |
| Walking | 95-percentile of walking cadence | 0.271 (0.038, 0.481) | No |
| Walking | Ambulatory minutes | -0.083 (-0.348, 0.154) | No |
| Walking | No. of walking bouts | -0.192 (-0.449, 0.052) | No |
| Walking | No. of long walking bouts-1min | -0.175 (-0.528, 0.078) | No |
| Walking | No. of long walking bouts-2min | 0.059 (-0.253, 0.28) | No |
| Walking | No. of short walking bouts-30sec-min | -0.186 (-0.438, 0.05) | No |
| Walking | No. of long walking bouts-5min | 0.33 (0.13, 0.499) | Yes |
| Walking | Step Count | 0.025 (-0.251, 0.25) | No |
| Walking | Peak 15-min cadence | -0.031 (-0.336, 0.207) | No |
| Walking | Peak 30-min cadence | 0.075 (-0.196, 0.327) | No |
| Walking | Peak 60-min cadence | 0.295 (0.055, 0.534) | Yes |
| Walking | Fraction of walking bouts-2min | 0.092 (-0.163, 0.369) | No |
| Walking | Fraction of walking bouts-5min | 0.158 (-0.108, 0.382) | No |
| Non-walking | 95-percentile of NWB duration | 0.527 (0.291, 0.825) | Yes |
| Non-walking | Mean NWB duration | 0.529 (0.347, 0.704) | Yes |
| Non-walking | Non-walking minutes | 0.073 (-0.182, 0.339) | No |
| Non-walking | No. of NWBs | -0.208 (-0.468, 0.037) | No |
| Non-walking | No. of long NWB-15min | -0.574 (-0.868, -0.331) | Yes |
| Non-walking | No. of long NWB-1min | -0.259 (-0.512, -0.018) | No |
| Non-walking | No. of long NWB-2min | -0.304 (-0.552, -0.064) | Yes |
| Non-walking | No. of short NWBs-30sec-min | -0.025 (-0.341, 0.225) | No |
| Non-walking | No. of long NWB-5min | -0.387 (-0.649, -0.135) | Yes |
| Composite | CM1 | 0.31 (0.09, 0.522) | Yes |
| Composite | CM2 | 0.428 (0.226, 0.629) | Yes |
| Composite | CM3 | -0.428 (-0.666, -0.21) | Yes |
| Composite | CM4 | 0.438 (0.218, 0.676) | Yes |
| Composite | CM5 | 0.138 (-0.102, 0.377) | No |
| Composite | CM6 | -0.098 (-0.383, 0.149) | No |
| Clinic-based | MDS-UPDRS Part 1+2+3 total (Off-State) | 0.24 | |
| Clinic-based | MDS-UPDRS Part 2+3 total (Off-State) | 0.29 | |
| Clinic-based | MDS-UPDRS Part 3 total (Off-State) | 0.11 | |
| Clinic-based | MDS-UPDRS Part 2 total | 0.09 | |
| Clinic-based | MDS-UPDRS Part 1+2+3 total (On-State) | 0.21 | |
| Clinic-based | MDS-UPDRS Part 2+3 total (On-State) | 0.16 | |
| Clinic-based | MDS-UPDRS Part 3 total (On-State) | 0.22 | |
